# Mood computational mechanisms underlying increased risk behavior in adolescent suicidal patients

**DOI:** 10.1101/2023.10.31.23297870

**Authors:** Zhihao Wang, Tian Nan, Fengmei Lu, Yue Yu, Xiao Cai, Zongling He, Yuejia Luo, Ting Wang, Bastien Blain

**Author notes:** Corresponding authors: Zongling He, M.D., Ph.D. The Clinical Hospital of Chengdu Brain Science Institute, School of Life Science and Technology, University of Electronic Science and Technology of China, Chengdu, 611731, PR China; Ting Wang, Ph.D. Institute for brain research and rehabilitation, South China Normal University, Guangzhou, 510631, PR China. These authors contributed equally to this work (Zhihao Wang, Tian Nan).

## Abstract

Suicidal thoughts and behaviors (STB) are among the leading causes of death worldwide. Although previous research has consistently documented elevated risk-taking in individuals with STB and identified mood disturbances as central features of suicidality, the precise cognitive and affective computational mechanisms underlying this increased risky behavior remain poorly understood. Here, 83 adolescent inpatients with affective disorders—including 58 patients with STB (S^+^) and 25 without STB (S^−^)—and 118 age-and sex-matched healthy controls (HC) completed a decision-making task involving choices between certain and gamble options, alongside momentary mood ratings. Behavioral analyses showed that S^+^ exhibited greater risk-taking than both S^−^ and HC. Computational modeling of choice behavior using a prospect-theory framework augmented with value-insensitive approach–avoidance parameters indicated that this increase in risky behavior was specifically driven by an elevated approach parameter in S^+^. In addition, mood-model analyses revealed reduced sensitivity to certain rewards in S^+^ relative to S^−^ and HC. Importantly, these computational signatures predicted suicidal symptom severity and showed generalizability in an independent general-population sample (n = 747). In S^+^, lower mood sensitivity to certain rewards was associated with greater gambling, providing a computational affective account of increased risk-taking in STB. These findings remained robust after adjusting for demographic, clinical, and medication-related variables. Overall, our study identifies cognitive and affective computational mechanisms contributing to elevated risk-taking in STB and highlights their potential relevance for the early identification and prevention of suicidality.

## Introduction

Every 40 seconds, a life is lost due to suicide(1). Suicidal thoughts and behaviors (STB) are one of leading causes of death worldwide that have devastating impacts on individuals, families, and societies. STB occurs from adolescence(2,3), especially in the context of mood disorders, e.g., major depressive disorder (MDD), anxiety disorder (AD), and bipolar disorder (BD)(4). Despite the progress made during the last 50 years for identifying risk factors (5) and developing preventing strategies(6), death rate from STB has not declined(7). The limited comprehension of cognitive and affective mechanisms creates a substantial gap in pinpointing targets for early prediction, screening, detection, and intervention in cases of suicidal thoughts. Understanding what is impaired in STB patients’ decision process would be key to prevent STB, for example through cognitive behavioral therapy (9,10).

Although meta-analyses have shown increased risk behavior in patients with STB across different risk domains (for a short summary, see Table S1) (11–13), the underlying cognitive computational mechanism is still unknown. Specifically, some studies found heightened loss aversion in STB in the context of the balloon analog risk task and the gambling task (14,15), while others observed the opposite pattern using the Iowa gambling task (16). Although all these results aligned with their hypotheses (for a short summary, see Table S1), this contradictory evidence may originate from the use of underspecified models. A growing literature indeed shows that risky behavior can be far better explained after adding value-insensitive approach and avoidance components to prospect theory(17,18), that is by including a decision bias in favor of the highest gain (approach) and another decision bias against the lowest loss (avoidance), above and beyond options value difference. This class of models highlights the important role of value-insensitive motivational components in decision making in addition to risk attitude-driven valuation (e.g., loss/risk aversion)(19). Importantly, STB has been proposed in theoretical work to result from abnormal motivational system(20–23), but no direct evidence support such proposals. Therefore, investigating motivational components may facilitate understanding why STB is associated with increased risk-taking behavior. We therefore hypothesized that heightened approach motivation, or weakened avoidance motivation, would account for increased risk behavior in STB.

While suicide is a decision process per se, atypical mood dynamics have been thought to be at the core of STB(3). Contemporary theories of suicide converge on the idea that STB is initially caused by low mood experience. The interpersonal theory of suicide proposes that suicidal desire arises when people simultaneously feel socially disconnected (“thwarted belongingness”) and like a burden on others (“perceived burdensomeness”), experiences that are tightly linked to chronically low mood(24). The motivational–volitional model(25) and the three-step theory (27,28) similarly emphasize that when negative mood and feelings of defeat or entrapment are experienced as inescapable, they can give rise to suicidal ideation, and that the progression from ideation to suicide attempts depends on additional factors such as reduced fear of death, increased pain tolerance, and a tendency to act impulsively under intense affect. Some official organizations, e.g., National Institute of Mental Health, have also listed mood problems as warning signals(8). Interestingly, within the framework of decision making under uncertainty, gambling on lotteries with a revealed outcome has been found to induce high mood variance(28), providing an opportunity to assess the relationship between deficient mood and increased gambling decisions in STB. Specifically, in a gambling task with momentary mood ratings (also referred to happiness or subjective well-being), where participants were asked to make decisions between certain vs. gamble options (2 possible outcomes, 50% probability for each), Rutledge et.al., (2014) found that mood was sensitive to certain rewards (CR), reward expectation (EV), and reward prediction error (RPE; the difference between experienced and expected outcome)(28). Although mood is thought to persist for hours, days, or even weeks(29–32), momentary mood, measured over the timescale in the laboratory setting, represents the accumulation of the impact of multiple events at the scale of minutes (29,31,33–37). Momentary mood external validity is demonstrated e.g., through its association with depression symptoms (36). Mood is different from emotions, which reflect immediate affective reactivity and is more transient (e.g. from surprise to fear) (30–32,38). Here, we investigated which mood computational components (among CR, EV and RPE) are associated with STB. We expect the mood response to gambling-related quantities (EV and RPE) to be higher in STB compared to the control groups. In contrast, riskier decisions may result from aversion to CR in STB. Therefore, another possibility is that lower mood sensitivity to CR would relate to increased risk behavior in STB.

To summarize, the aim of this study is to examine cognitive and affective computational mechanisms underlying increased risk behavior in adolescent patients with STB, as adolescent period might provide a developmental window for opportunities for early intervention(2). This study aligns with the principles of Computational Psychiatry(39), which assumes that psychiatric symptoms arise from alterations in cognitive and affective computations. The ultimate aim for this field is to uncover ‘‘computational phenotypes’’ — distinct patterns of computational dysfunctions — potentially enabling targeted treatments, improved outcome predictions, and more precise diagnostic frameworks. Specifically, we employed a gambling task with momentary mood ratings to assess risk behavior and track mood fluctuations in response to various events. We applied computational models of risky decision making and momentary mood to dissect cognitive and affective processes contributing to heightened risky behavior in patients with STB. Regarding choices, we hypothesized heightened approach motivation, or weakened avoidance motivation, in STB, which would account for increased risk behavior. Regarding mood dynamics, we hypothesized that greater mood sensitivity to gambling-related variables (i.e., RPE and EV), or reduced mood sensitivity to CR, would explain increased risk behavior in STB.

## Methods and materials

### Participants

We recruited 95 adolescent patients with mood disorder from the Clinical Hospital of Chengdu Brain Science Institute, University of Electronic Science and Technology of China (The Mental Health Center of Chengdu, Sichuan, China). According to medical records and information from family and friends by the researcher (T.N) and psychiatrists (F.L, Y.Y, X.C, & Z.H), patients with suicidal thoughts and behaviors were categorized as suicidal group (S^+^), while patients without suicidal thoughts and behaviors were identified as control group (S^-^). The definition for suicidal thoughts in this study was active thoughts of suicide, i.e., wishing to die and having some intention to do so (see Supplementary Note 1 for details). This grouping operation was consistent with previous suicidal-related literature (40–45), reflecting the general tendency for suicidal risks among adolescence. As baseline control, we also recruited 124 sex- and age-matched healthy adolescents (HC). We assert that all procedures contributing to this work comply with the ethical standards of the ethical committee of The Clinical Hospital of Chengdu Brain Science Institute, University of Electronic Science and Technology of China (number: 2022(33)) on human experimentation and with the Helsinki Declaration of 1975, as revised in 2008. All procedures involving human subjects/patients were approved by the ethical committee of The Clinical Hospital of Chengdu Brain Science Institute, University of Electronic Science and Technology of China (number: 2022(33)). Informed written consent was obtained. Patients were included if they met the following criteria: 1) both the researcher and psychiatrists agreed on their group classification; 2) they had a current diagnosis of major depressive disorder (MDD; unipolar depression), generalized anxiety disorder (GAD), or bipolar disorder with depressive episodes (BD), confirmed by two experienced psychiatrists using the Structured Clinical Interview for DSM-IV-TR-Patient Edition (SCID-P, 2/2001 revision; see Supplementary Note 1 for details); 3) they were between 10 and 19 years of age; 4) they had no organic brain disorders, intellectual disability, or head trauma; 5) they had no history of substance abuse; 6) they had no experience of electroconvulsive therapy. In addition, participants were excluded if they failed more than 1/4 of the catch trials. The final sample consisted of 25 patients for S^-^, 58 patients for S^+^, and 118 HC participants. See Table 1 and Table S2 for demographic, clinical and psychological information. The validation dataset was from our previous online study, with 747 general participants completing the same task and numerous anxiety/depression-related questionnaires for different purposes. See (46) for demographic and psychological details.

**Table 1.**
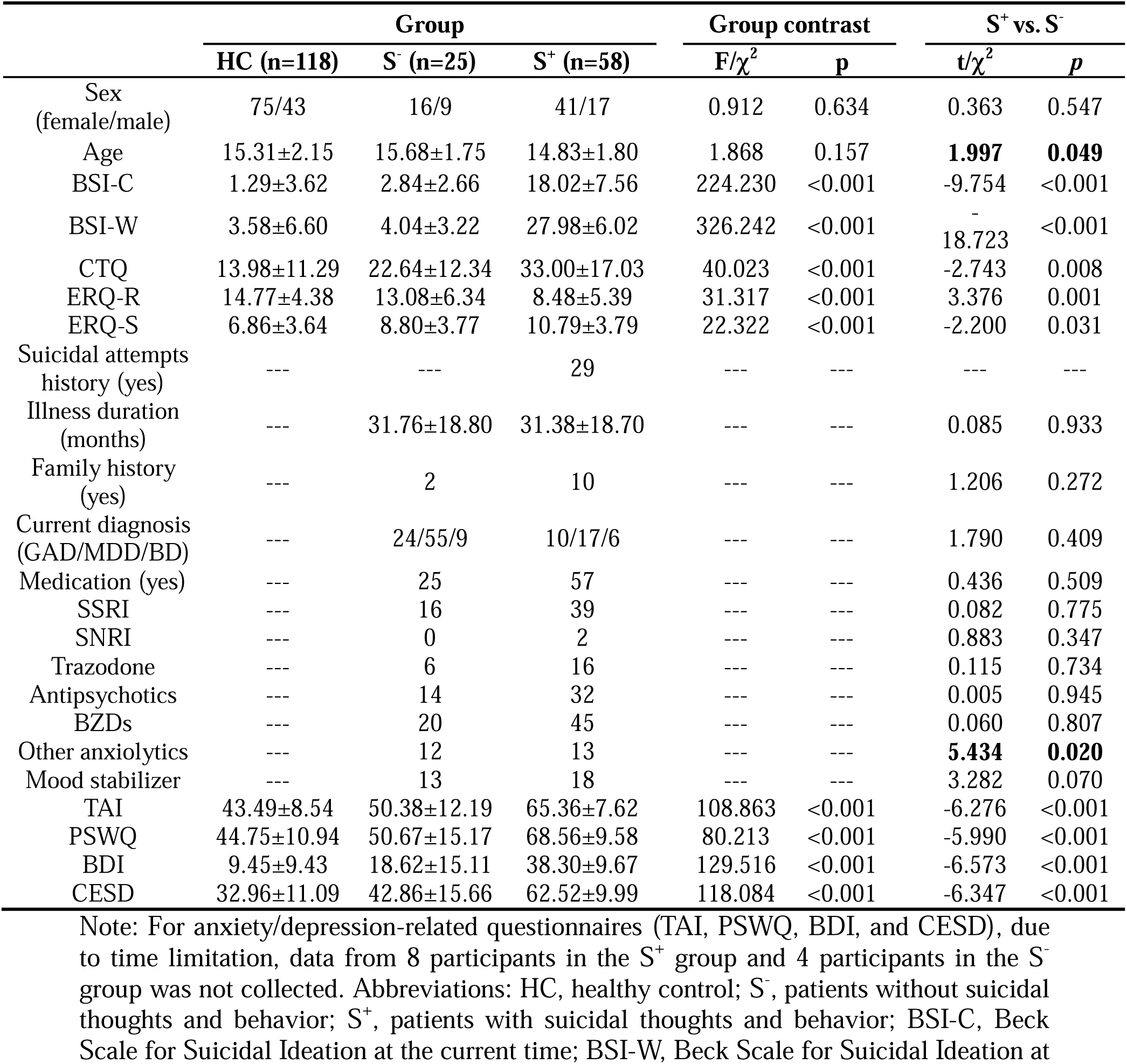
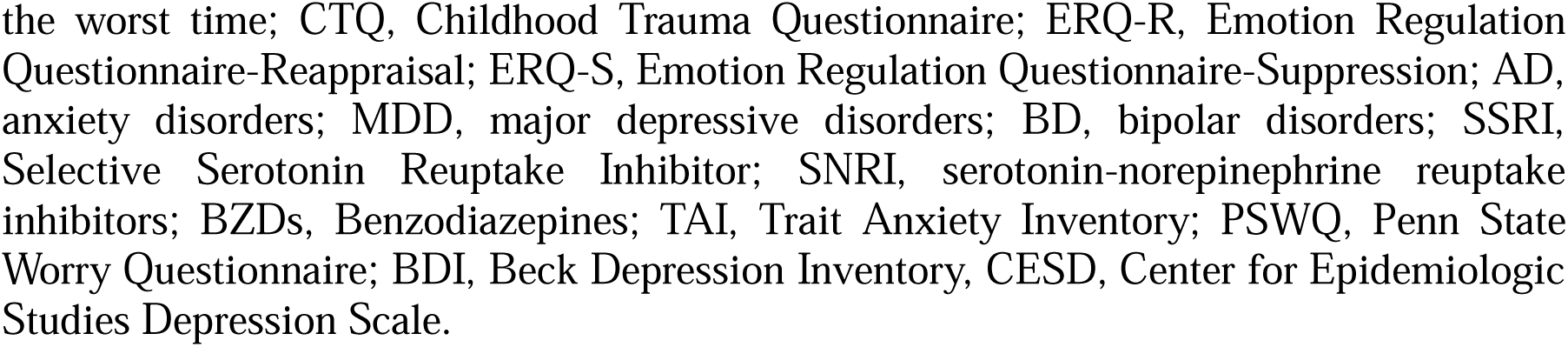
Demographics, clinical, psychological characteristics of patients with and without suicidal thoughts and behaviors.

|  | Group |  |  | Group contrast |  | S<sup>+</sup> vs. S<sup>-</sup> |  |
| --- | --- | --- | --- | --- | --- | --- | --- |
| | HC (n=118) | S<sup>-</sup> (n=25) | S<sup>+</sup> (n=58) | F/$\chi^2$ | p | t/$\chi^2$ | p |
| Sex<br>(female/male) | 75/43 | 16/9 | 41/17 | 0.912 | 0.634 | 0.363 | 0.547 |
| Age | 15.31±2.15 | 15.68±1.75 | 14.83±1.80 | 1.868 | 0.157 | **1.997** | **0.049** |
| BSI-C | 1.29±3.62 | 2.84±2.66 | 18.02±7.56 | 224.230 | <0.001 | -9.754 | <0.001 |
| BSI-W | 3.58±6.60 | 4.04±3.22 | 27.98±6.02 | 326.242 | <0.001 | -18.723 | <0.001 |
| CTQ | 13.98±11.29 | 22.64±12.34 | 33.00±17.03 | 40.023 | <0.001 | -2.743 | 0.008 |
| ERQ-R | 14.77±4.38 | 13.08±6.34 | 8.48±5.39 | 31.317 | <0.001 | 3.376 | 0.001 |
| ERQ-S | 6.86±3.64 | 8.80±3.77 | 10.79±3.79 | 22.322 | <0.001 | -2.200 | 0.031 |
| Suicidal attempts<br>history (yes) | --- | --- | 29 | --- | --- | --- | --- |
| Illness duration<br>(months) | --- | 31.76±18.80 | 31.38±18.70 | --- | --- | 0.085 | 0.933 |
| Family history<br>(yes) | --- | 2 | 10 | --- | --- | 1.206 | 0.272 |
| Current diagnosis<br>(GAD/MDD/BD) | --- | 24/55/9 | 10/17/6 | --- | --- | 1.790 | 0.409 |
| Medication (yes) | --- | 25 | 57 | --- | --- | 0.436 | 0.509 |
| SSRI | --- | 16 | 39 | --- | --- | 0.082 | 0.775 |
| SNRI | --- | 0 | 2 | --- | --- | 0.883 | 0.347 |
| Trazodone | --- | 6 | 16 | --- | --- | 0.115 | 0.734 |
| Antipsychotics | --- | 14 | 32 | --- | --- | 0.005 | 0.945 |
| BZDs | --- | 20 | 45 | --- | --- | 0.060 | 0.807 |
| Other anxiolytics | --- | 12 | 13 | --- | --- | **5.434** | **0.020** |
| Mood stabilizer | --- | 13 | 18 | --- | --- | 3.282 | 0.070 |
| TAI | 43.49±8.54 | 50.38±12.19 | 65.36±7.62 | 108.863 | <0.001 | -6.276 | <0.001 |
| PSWQ | 44.75±10.94 | 50.67±15.17 | 68.56±9.58 | 80.213 | <0.001 | -5.990 | <0.001 |
| BDI | 9.45±9.43 | 18.62±15.11 | 38.30±9.67 | 129.516 | <0.001 | -6.573 | <0.001 |
| CESD | 32.96±11.09 | 42.86±15.66 | 62.52±9.99 | 118.084 | <0.001 | -6.347 | <0.001 |

Note: For anxiety/depression-related questionnaires (TAI, PSWQ, BDI, and CESD), due to time limitation, data from 8 participants in the S<sup>+</sup> group and 4 participants in the S<sup>-</sup> group was not collected. Abbreviations: HC, healthy control; S<sup>-</sup>, patients without suicidal thoughts and behavior; S<sup>+</sup>, patients with suicidal thoughts and behavior; BSI-C, Beck Scale for Suicidal Ideation at the current time; BSI-W, Beck Scale for Suicidal Ideation at
the worst time; CTQ, Childhood Trauma Questionnaire; ERQ-R, Emotion Regulation Questionnaire-Reappraisal; ERQ-S, Emotion Regulation Questionnaire-Suppression; AD, anxiety disorders; MDD, major depressive disorders; BD, bipolar disorders; SSRI, Selective Serotonin Reuptake Inhibitor; SNRI, serotonin-norepinephrine reuptake inhibitors; BZDs, Benzodiazepines; TAI, Trait Anxiety Inventory; PSWQ, Penn State Worry Questionnaire; BDI, Beck Depression Inventory, CESD, Center for Epidemiologic Studies Depression Scale.

### Self-reported questionnaires

Participants completed a set of Chinese-version suicidal-, emotion regulation-, and depression/anxiety-related questionnaires. These measurements included the Beck Scale for Suicidal Ideation at the current time (BSI-C, 19 items) and at the worst time (BSI-W, 19 items)(47), the Childhood Trauma Questionnaire (CTQ, 28 items)(48), Emotion Regulation Questionnaire-Reappraisal (ERQ-R, 6 items) and Suppression (ERQ-S, 4 items)(49). In addition, as patients were available only for a limited duration, anxiety/depression-related scales from only 50 participants in the S^+^ group and only 21 participants in the S^-^ group were collected. Specifically, patients filled the Trait subscale of the State-Trait Anxiety Inventory (TAI; 20 items)(50), the Penn State Worry Questionnaire (PSWQ; 16 items), the Beck Depression Inventory (BDI; 21 items)(51), and the Center for Epidemiologic Studies Depression Scale (CESD; 20 items)(52).

### Experimental Procedure

Participants were asked to make a choice between a certain option and a gamble (50% probability for each outcome) to maximize their points and to rate their momentary moods(17,28). Before performing the task, participants were asked to rate their current happiness that we consider as their initial mood. At the beginning of the task, participants were endowed with 500 points. Each trial started with two options (a gamble option and a certain option) which were presented randomly on each side (Figure 1A). Upon response, the chosen option was highlighted in yellow for 0.5 s. Note that Rutledge et al., (2014) displayed the chosen option for about 6 s(28), a delay we shortened for the sake of time. Then the corresponding outcome at the screen center was presented for 1 s, followed by a fixation cross with a random duration (0.6∼1.4 s). If the gamble was chosen, participants had equal probability to obtain each outcome. The obtained outcome was added to their total score, which was presented at the top-right corner. Every 2∼3 trials, participants rated their mood (“how happy are you at this moment?”) from 0 (very unhappy) to 100 (very happy) by moving a slider anchored at midpoint (i.e., 50). Upon identifying their current mood, a fixation cross was presented with a random duration (0.6∼1.4 s). This task consisted of 90 randomly presented trials, including 30 mixed trials, 30 gain trials, and 30 loss trials. The numbers of choice trials and mood ratings were comparable to those in prior computational modeling studies (34,35). In mixed trials, participants made a choice between a certain amount 0 and a gamble with a gain amount {40, 45, or 75} and a loss amount determined by a multiplier {0.2, 0.34, 0.5, 0.64, 0.77, 0.89, 1, 1.1, 1.35, or 2} on the gain amount. For example, with a gain amount of 40 and a multiplier of 2 for the loss (2 times 40 = 80), participants chose between a certain option of 0 and a gambling option, which offered a 50% chance to win 40 and a 50% chance to lose 80. These trials are therefore particularly suited to measuring loss aversion. In gain trials, there was a certain gain amount {35, 45, or 55} and a gamble with 0 and a gain amount determined by a multiplier {1.68, 1.82, 2, 2.22, 2.48, 2.8, 3.16, 3.6, 4.2, or 5} on the certain gain amount. In loss trials, there were a certain loss amount {-35, -45, or -55} and a gamble with 0 and a loss amount determined by a multiplier {1.68, 1.82, 2, 2.22, 2.48, 2.8, 3.16, 3.6, 4.2, or 5} on the certain loss amount. We also added an extra 4 trials in the entire task for attentional checks. For example, participants were asked to make a choice between a certain gain 20 and a gambling 35/55, where the correct response for this trial was the gambling choice (as the worst lottery outcome was higher than the certain reward). All experimental procedures were programmed using Psychopy3 (2021.2.3).

**Figure 1.**
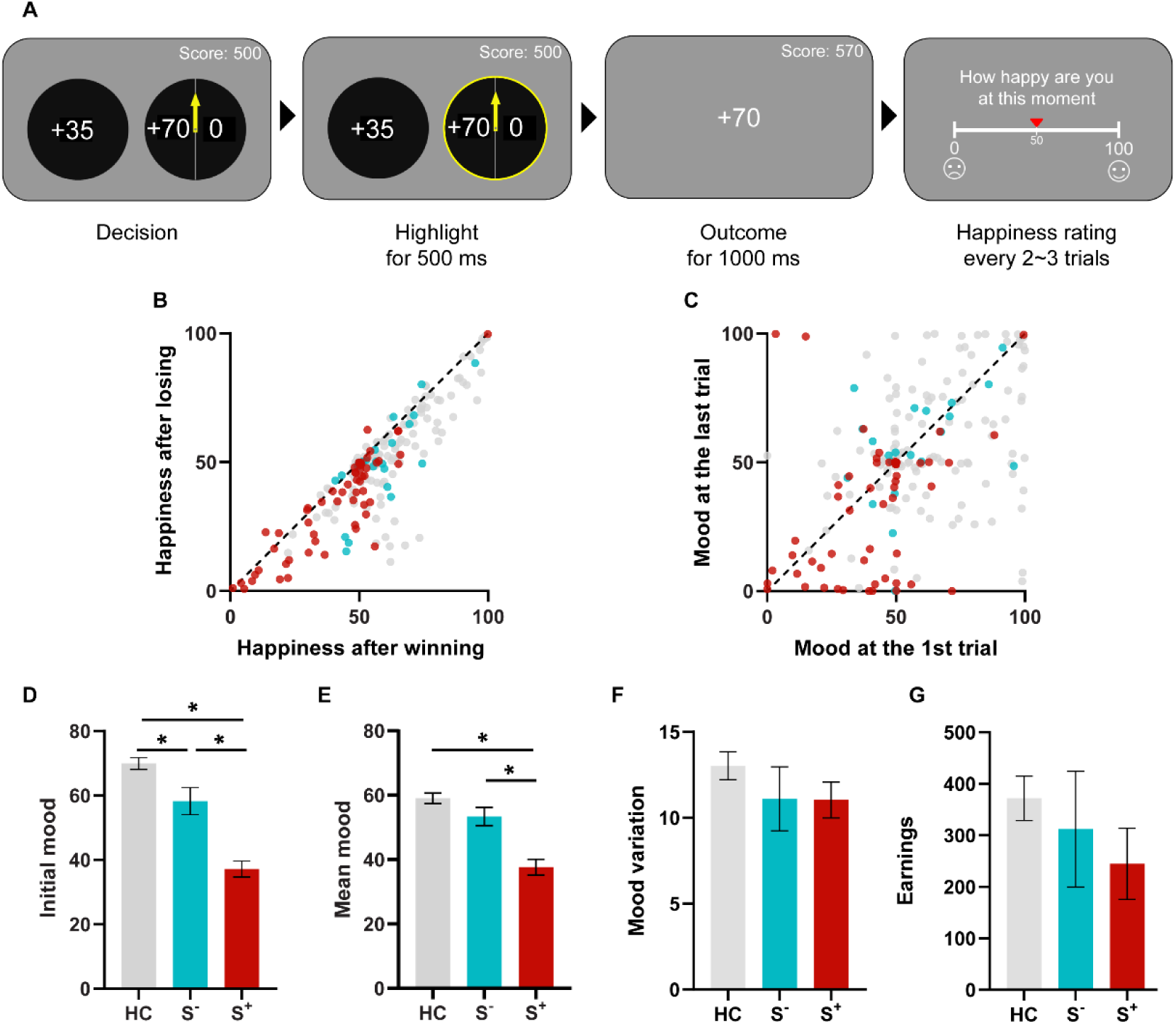
Task design, outcome and time effects on mood, and group differences in mood. A) Gambling task with mood ratings. On each trial, participants were asked to choose between a certain option and a gambling option (self-paced). Once selected, the chosen option was highlighted in yellow for 500 ms. Then the corresponding outcome was displayed in the center of the screen for 1000 ms. The cumulated score was always shown in the right-upper corner. Every 2 to 3 trials, participants were asked to complete a self-paced rating of their happiness, answering the question “How happy are you at the moment” on a slider from 0 (very unhappy) to 100 (very happy). B) Patients and healthy controls felt happier after winning than losing. C) Mood drifted over time. D) Group difference in mood before the task shows weakened mood in S^+^. E) Group difference in average mood displays lower mood experience in S^+^. F) Mood variance was similar for all the three groups, as indexed by standard deviation of happiness ratings across the task. G) Each group earned about the same amount of points by the end of the task. Abbreviations: HC, healthy control; S^-^, patients without suicidal thoughts and behavior; S^+^, patients with suicidal thoughts and behavior; \**p*<0.05. Error bars correspond to the standard error.

### Choice computational models

In line with previous studies(17,18), our choice model space included expected value model (cM1), prospect theory model (cM2)(53), and approach-avoidance prospect theory model (cM3)(17). For cM2 (Equations 1-4), there were 3 parameters, including risk aversion (α, range: [0.3, 1.3]), loss aversion (λ: [0.5, 5]), and inverse temperature (μ: [0, 10]).

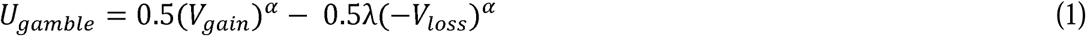

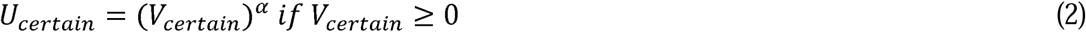

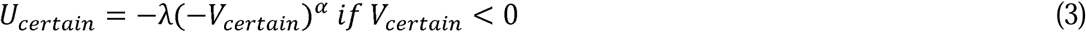

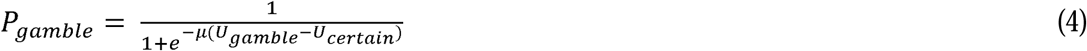

where V*_gain_* and V*_loss_* are the objective gain and loss from a gamble, respectively. Please note that V*_gain_*is 0 in loss trials and V*_loss_* is 0 in gain trials. V*_certain_*is the objective value for the certain option. U*_gamble_* and U*_certain_* denote the subjective utilities of the gamble and the certain option, respectively. Choice probability for gamble (P*_gamble_*) is determined by the softmax rule. Building on cM2, cM3 decomposes the decision process into risk-attitude-driven valuation (e.g., loss and risk aversion) and value-insensitive motivational components (Equations 1-3 & 5-7). That is, choice probability for P*_gamble_* in cM3 is jointly determined by the softmax rule and approach/avoidance parameters (*β_gain_*: [-1, 1], *β_loss_*: [-1, 1]). Approach/avoidance parameters are not applied in mixed trials. Please note that a higher gambling rate does not imply a change in risk attitude per se: it can arise from an increased value-insensitive approach bias even when risk-attitude parameters are comparable between groups. Risk attitude is indeed conceptualized in economics as the curvature of the utility function (i.e., the subjective value) of the objective outcomes, with concave curves associated with risk aversion, and convex curves associated with risk seeking (54,55). By contrast, the approach or avoidance bias apply to all the value. A possible interpretation of the approach bias is that participant approach the option with the highest possible gain (the lottery) in the gain frame; the avoidance bias would then reflect a tendency to systematically avoid the highest potential losses (the lottery) in the loss frame.

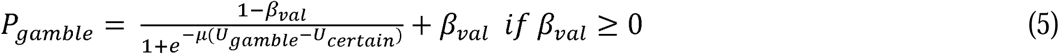

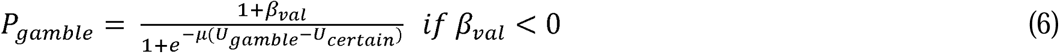

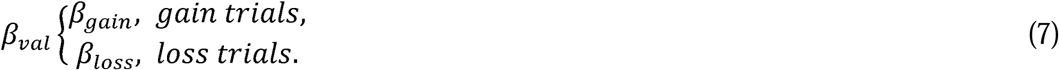

### Mood computational models

To quantify how different events impacted participants’ momentary mood during the gambling task, we conducted a stage-wise model construction procedure(56). That is, we added or removed each component to the model progressively, based on the best model from the previous stage. In Stage 1, we fit the classic model assuming that momentary mood depends on the recency-weighted average of the chosen certain reward (CR), expected value of the chosen gamble (EV), and reward prediction error (RPE; mM1; Equation 8). RPE was defined as the difference between the obtained and expected value.

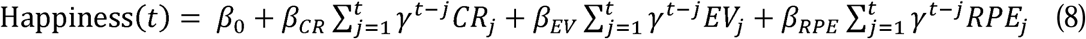

Here, *t* and *j* are trial numbers, *β*_0_ is a baseline mood parameter, other weights *β* capture the influence of different event types, γ ɛ [0,1] is a decay parameter representing how many previous trials influence happiness. CR*_j_* is the CR if the certain option was chosen on trial *j*; otherwise, CR*_j_* is 0. EV*j* is the EV and RPEj is the RPE on trial *j* if the gamble was chosen. If the certain option was chosen, then EV*_j_* = 0 and RPE*j* =0.

To check that mood ratings are best explained by a shared forgetting factor (i.e., the recency-weighted history of different event types), we compared a model with a single decay parameter to an alternative model, including forgetting factors for each event type, e.g., different decay parameters for CR, EV, and RPE (mM2; Equation 9).

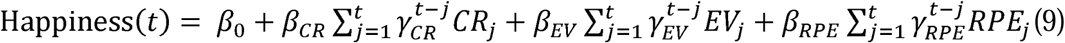

In Stage 2, to identify whether mood can be better explained by RPE, we fit an alternative model in which mood ratings are explained by the recency-weighted average of the certain reward (CR) and the gamble reward (GR; mM3; Equation 10), a simple model providing a mood sensitivity parameter for certain rewards and gamble rewards. We also fit a model with two forgetting factors, one for CR and one for GR (mM4; Equation 11).

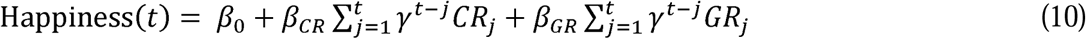

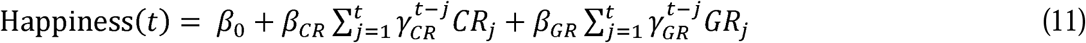

In Stage 3, to check whether mood data can be better explained by a single event (CR or GR), we compared a CR-mood model (mM5) and a GR-mood model (mM6).

### Model fitting and comparison

We fit model parameters by using the method of maximum likelihood estimation (MLE) with fmincon function of MATLAB (version R2015a) at the individual level. To avoid local minimum, we ran this optimization function with random starting locations 50 times. Bayesian information criteria (BIC) were used to compare model fits.

### Replication of suicidal-related results in an independent dataset (n = 747)

We next verified our results in an independent dataset, including the same task and BDI questionnaire in 747 general participants (500 females; age: 20.90±2.41) (46). One item in BDI involves the measurement of STB. In item 9 of BDI, participants chose one option that describes them best: Option 1, “I don’t have any thoughts of killing myself.”; Option 2, “I have thoughts of killing myself, but I would not carry them out.”; Option 3, “I would like to kill myself.”; Option 4, “I would kill myself if I had the chance.”. In line with the current definition of S^+^/S^-^ in the clinical dataset, we identified S^+^ group as choosing Option 2, 3, or 4, while participants selecting Option 1 were categorized as S^-^group. Therefore, there were 129 participants in S^+^ and 618 participants in S^-^. We did not find significant group difference in sex and age (*ps* > 0.075). To make it comparable, we fit the winning choice and mood models from the clinical study.

## Predictive model of suicidal risks

### Internal Validation

To evaluate the out-of-sample predictive utility of computational parameters for STB, we used lasso regression within a repeated nested 5-fold cross-validation framework. In each of 100 iterations, the full sample was randomly divided into five folds. For each outer fold, the model was trained on four folds and tested on the remaining fold. Predictor variables were z-scored within the training data, and the corresponding training-set means and standard deviations were then applied to normalize the test data. Within each training set, the lasso penalty parameter was selected via an inner 5-fold cross-validation procedure using the minimum mean squared error criterion. The resulting coefficients were then used to generate predictions for the held-out fold. After all outer folds had been completed, the cross-validated predictions for all participants were combined, and model performance was quantified as the Spearman correlation between predicted and observed STB scores, given that suicidal symptom scores (BSI-C) were not normally distributed (Kolmogorov–Smirnov test, *p* < 0.001). This entire procedure was repeated 100 times to obtain a stable estimate of predictive performance.

### External Validation

To further assess robustness and generalizability beyond the original sample, we conducted an external validation analysis using an independent dataset (*n* = 747). Specifically, regression coefficients and intercepts were averaged across folds and repetitions from the internal validation procedure to derive a stable final model. This model was then applied to the external dataset, using the same predictors (β_gain_ and β_CR_), to generate predicted scores. External validity was assessed by calculating the Spearman correlation between model-predicted scores and scores on item 9 of BDI.

### Statistical analysis

We performed chi-square, independent-sample t-test or repeated measure ANOVA to test group-related differences. Spearman correlations were used to check correlations among suicidal-related questionnaires, choice data, and mood data. Generalized linear model was conducted for control analysis using Matlab R2015a. Mediation analysis was conducted using R (4.1.0) and the R package ‘mediation’. All reported tests are two-tailed unless otherwise specified. For the replication of previous findings in the validation dataset, we used one-tailed tests in line with our clinically motivated directional hypothesis. We set the significance level at p = 0.05. Multiple comparisons were corrected using Benjamini-Hochberg false discovery rate (FDR) correction (see Supplementary Note 8 for details).

## Results

### Demographic and clinical characteristics

Overall, sex and age were comparable among S^+^, S^-^, and HC groups (*ps* > 0.157), though S^+^ was significantly younger than S^-^ (t = 1.997, *p* = 0.049). As expected, S^+^ scored significantly higher than S^-^ and HC in suicidal-related scales (e.g., BSI-C; *ps* < 0.001), further validating our grouping of participants. There was no significant difference between S^+^ and S^-^ in illness duration, family history, diagnosis, and various medications use (*ps* > 0.07), except other anxiolytics (χ^2^=5.434, *p* = 0.020). See Table 1 and Table S2 for details.

### Sanity checks

To ensure engagement and task validation, we performed sanity checks. As expected, we found significant group differences in psychological measurements (*ps* < 0.001), including childhood trauma, emotion regulation, and anxiety/depression (Table 1 and Table S2). In addition, we replicated the classic mood-related effects(57,58): 1) subjects were happier after winning than losing (t = 11.001, *p* < 0.001; Figure 1B) and 2) mood drifted over time (t = -3.254, *p* = 0.001; Figure 1C). As grouping checks, we found a hierarchical pattern of mood level both before the task and across the task (S^+^ < S^-^ < HC; for initial mood, F = 53.415, *p* < 0.001; S^+^ vs. S^-^: t = -4.525, *p* < 0.001; S^+^ vs. HC: t = - 10.427, *p* < 0.001; S^-^ vs. HC: t = -2.634, *p* = 0.009; Figure 1D; for mean mood, F = 28.018, *p* < 0.001; S^+^ vs. S^-^: t = -3.773, *p* < 0.001; S^+^ vs. HC: t = -7.292, *p* < 0.001; S^-^ vs. HC: t = -1.458, *p* = 0.147; Figure 1E). No significant group difference in mood variation was found (F = 1.270, *p* = 0.284; Figure 1F) which suggests that any parameter difference between groups is unlikely to be explained by mood variance. Moreover, there was no group difference in terms of mood drift effect, or earnings (Figure 1G; *ps* > 0.276).

### Patients with suicidal thought and behavior approached gambles more than patient controls and healthy controls, while risk attitude was comparable across groups

To replicate previous findings of increased risk behavior in suicidal populations, we conducted a two-way ANOVA on gambling rate with group (S^+^/S^-^/HC) as a between-subject factor and trial type (mix/gain/loss) as a within-subject factor. We found a significant main effect of group (F = 3.655, *p* = 0.028, partial *η*^2^ = 0.036; Figure 2A), with more gambling behavior for S^+^ than S^-^ (two-sample t-test, t = 2.145, *p* = 0.035) and HC (t = 2.465, *p* = 0.015) and comparable gambling behavior between S- and HC (t = - 0.439, *p* = 0.661) across the task. We also observed the main effect of trial type (F = 51.225, *p* < 0.001, partial *η*^2^ = 0.206; gain > mix > loss). We did not observe any significant interaction effect between group and trial type (F =0.270, partial *η*^2^ = 0.003). Within patients, this group effect on gambling rate remained significant after controlling for sex, illness duration, family history, diagnosis, and various medications use (*ps* < 0.05), as well as general symptoms (e.g., depression and anxiety; *p* = 0.024; also see Figure S4 and Table S10). Given high correlations among anxiety and depression questionnaires (rs > 0.753, *ps* < 0.001), we performed Principal Components Analysis (PCA) to extract main components, where each component explained 86.95%, 7.09%, 3.27%, and 2.68% variance, respectively. To further control for anxiety and depression, linear regression using these components as covariates revealed that the group effect on gambling rate remained significant (*p* = 0.024; Table S11). There was also no significant age/other anxiolytics use difference in gambling behavior (*ps* > 0.109; Figure S2).

**Figure 2.**
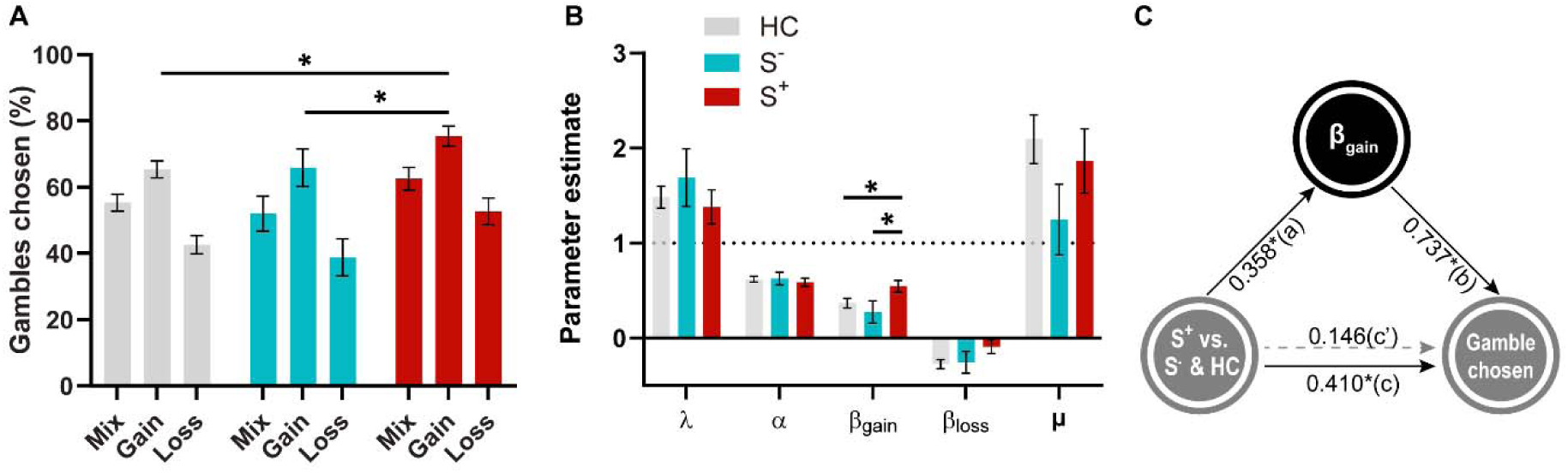
Choice results. A) Group differences in gambling behavior. The grey dots represent the winning model prediction. B) The estimated parameters from the winning choice model differed across groups. S^+^ exhibited stronger approach motivation than S^-^ and HC. C) The mediation model among the group, β*_gain_*, and gambling behavior in the gain condition. The approach parameter mediated the effects of STB group on increased gambling behavior in the gain condition. Abbreviations: HC, healthy control; S^-^, patients without suicidal thoughts and behavior; S^+^, patients with suicidal thoughts and behavior; \**p*<0.05.

We next performed a model comparison to select the model that best explains choice data. This analysis revealed that the winning model to formally quantify mechanisms for observed risky behavior is the approach-avoidance prospect theory model (cM3; mean R^2^ = 0.37; Table 2). Parameter and model recovery analyses showed that each model and parameter can be identified (see Supplementary Note 5; Figure S5 & S6). As predicted, we found a (marginally) significant group effect in approach parameter (F = 2.989, *p* = 0.053; Figure 2C), with a significant stronger approach motivation for S^+^ than S^-^ (t = 2.217, *p* = 0.029) and HC (t = 2.091, *p* = 0.038), and comparable between S- and HC (t = -0.737, *p* = 0.463). No other significant group difference in these parameters was found (*ps* > 0.135). Within patients, this group effect on the approach parameter remained significant after controlling for sex, illness duration, family history, diagnosis, and various medications use (*ps* < 0.05), as well as general symptoms (e.g., depression and anxiety; *p* = 0.027; also see Figure S4, Table S10). Linear regression using PCA components as covariates revealed that the group effect on approach parameter remained significant (*p* = 0.027; Table S11). There was also no significant age/other anxiolytics use difference in gambling behavior (*ps* > 0.223; Figure S2). Given significant correlations between group, approach parameter, and gambling rate for gain trials (*ps* < 0.017), we further conducted a mediation analysis with the assumption of the mediating effect of approach motivation of suicidality on the risk behavior. Given that we aimed to test the effect of STB, with S- and HC as controls, and S^−^ and given that HC did not differ in gambling behavior or in the approach parameter, we merged these two groups for the mediation analysis. Results supported our hypothesis (a×b = 0.321, 95% CI = [0.070, 0.549], *p* = 0.016; Figure 2C), confirming that suicidal thoughts and behavior increase risk behavior through stronger approach motivation.

**Table 2.**
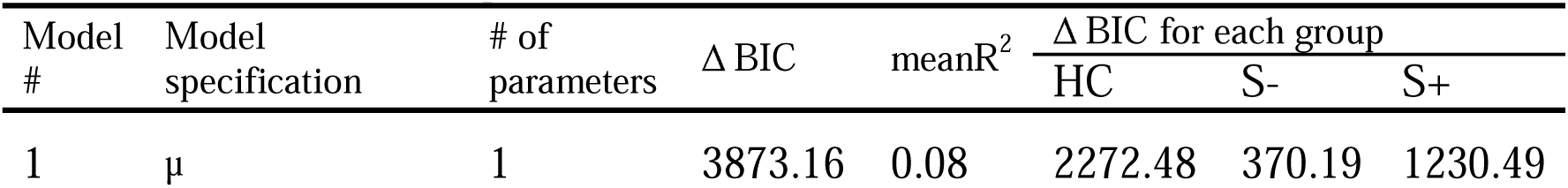
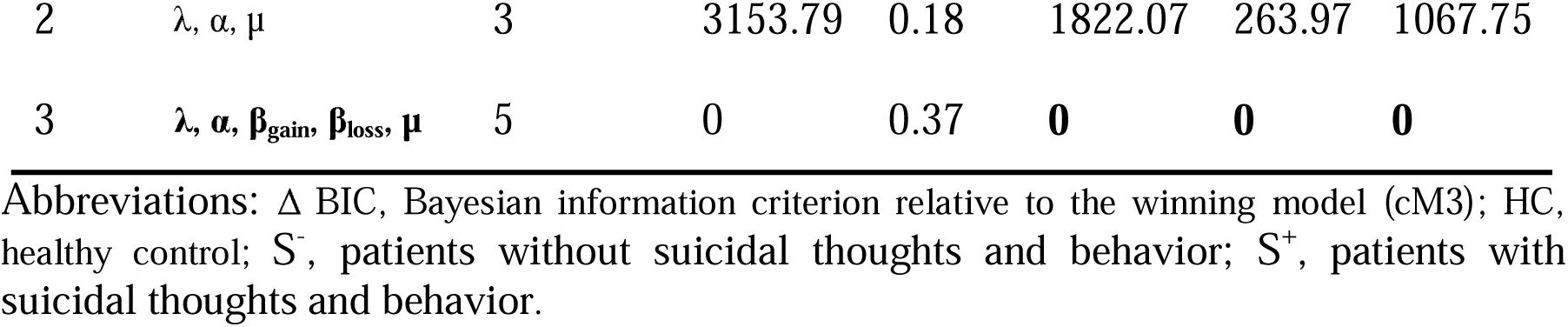
Choice model comparison.

### Mood sensitivity to certain rewards was reduced in patients with suicidal thought and behavior compared to patient controls and healthy controls

Next, we turned to mood model comparison. We observed inconsistent mood winning models for different groups (Table 3), suggesting an effect of STB on mood dynamics. Given that the focus of the current study was STB effect, especially for the S^+^ group, with the baseline control of S^-^ and HC groups, we specially focused on the winning model from the S^+^ group. Parameter and model recovery analyses showed that each model and parameter can be identified (see Supplementary Note 5; Figure S5 & S6 and TableS7). The winning mood model from S^+^ assumed that momentary mood fluctuations were explained by the recency-weighted average of certain reward (CR) and the gamble reward (GR; mM3; mean R^2^ = 0.42; Table 3). Overall, both CR and GR weights were significantly higher than 0 (CR: t = 8.033, *p* < 0.001; GR: t = 9.853, *p* < 0.001). The baseline parameter β_0_ was significant correlated with the initial mood (rho = 0.580, *p* < 0.001), validating this model. We also replicated previous depression-related findings (59): depression symptom measured by Beck Depression Inventory (BDI) was negatively correlated with the baseline mood parameter *β*_0_ (rho = -0.530, *p* < 0.001; Figure S7). We found significantly lower β_0_ in S^+^ than S^-^ (F = 22.861, *p* <0.001; t = -3.513, *p* < 0.001) and HC (t = -6.606, *p* < 0.001), which mirrors the lower initial mood pattern. Importantly, a two-way ANOVA on mood parameters with group (S^+^/S^-^/HC) as a between-subject factor, event type (CR/GR) as a within-subject factor showed a significant main effect of group (F = 3.835, *p* = 0.023, partial *η*^2^ = 0.037), with lower mood sensitivity for S^+^ than S^-^ (t = -2.080, *p* = 0.041) and HC (t = -2.758, *p* = 0.006) and comparable between S- and HC (t = -0.110, *p* = 0.913). We also observed a significant interaction effect between group and event type (F = 4.283, *p* = 0.015, partial *η*^2^ = 0.041; Figure 3B). Simple effect analysis revealed that S^+^ group exhibited significant lower mood sensitivity to CR as compared to GR (F = 4.823, *p* = 0.029, partial *η*^2^ = 0.024), while there was no significant CR-GR difference in S^-^ (although trendy; F = 2.783, *p* = 0.097, partial *η*^2^ = 0.014) and HC (F = 0.989, *p* = 0.321, partial *η*^2^ = 0.005). This interaction was driven by the group difference in CR (F = 6.085, *p* = 0.003, partial *η*^2^ = 0.058) rather than in GR (F = 0.801, *p* = 0.450, partial *η*^2^ = 0.008). Specifically, S^+^ showed lower mood sensitivity to CR than S^-^ (t = -2.661, *p* = 0.009) and HC (t = -3.381, *p* <0.001), while S^-^ and HC were comparable (t = 0.450, *p* = 0.679), suggesting S^+^ was specifically more insensitive to certain outcome than gamble outcome. No significant main event type (CR vs. GR) effect was found (F = 0.285, *p* = 0.594, partial *η*^2^ = 0.001). Within patients, this group effect on β_CR_ remained significant after controlling for gambling rate, earnings, mood-related outcome effect, mood drift effect, sex, illness duration, family history, diagnosis, and various medications use (*ps* < 0.032), as well as general symptoms (e.g., depression and anxiety; *p* = 0.001; also see Figure S4 and Table S10). Linear regression using PCA components as covariates revealed that the group effect on this mood parameter remained significant (*p* = 0.001; Table S11). There was also no significant age/other anxiolytics use difference in gambling behavior (ps > 0.582; Figure S2). These results indicate decreased mood sensitivity for certain rewards in suicidal populations.

**Figure 3.**
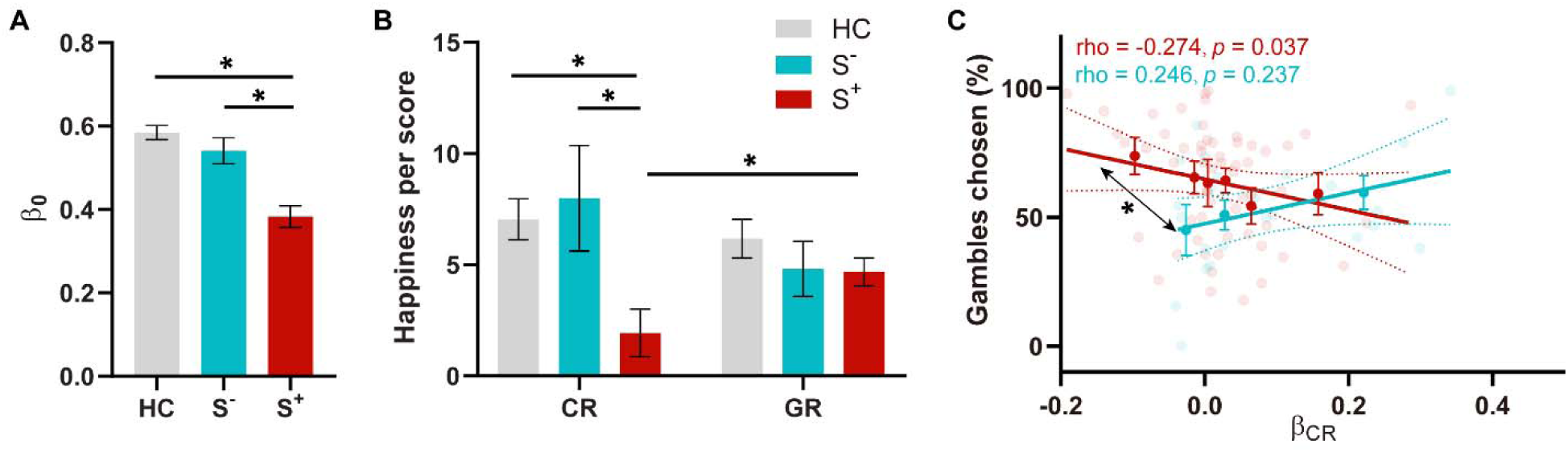
Effect of Suicidal thoughts and behavior on mood dynamics. A) Group difference in mood baseline, β_0_. B) Group differences in mood sensitivity to certain reward (CR) and gamble reward (GR). C) Correlational difference in S^-^ and S^+^ between mood sensitivity to CR and gambling behavior. The lighter, semi-transparent dots represent individual participants, while the dark dot with an error bar indicates the mean of binned scores (for illustration purposes only). Abbreviations: CR, certain reward; GR, gamble reward; HC, healthy control; S^-^, patients without suicidal thoughts and behavior; S^+^, patients with suicidal thoughts and behavior; \**p*<0.05.

**Table 3.** Mood model comparison.

| Model # | Model specification | # of parameters | $\Delta$ BIC | meanR<sup>2</sup> | $\Delta$ BIC for each group | | |
| --- | --- | --- | --- | --- | --- | --- | --- |
|  |  |  |  |  | HC | S<sup>-</sup> | S<sup>+</sup> |
| 1 | $\beta_0, \beta_{CR}, \beta_{EV}, \beta_{RPE}, \gamma$ | 5 | -106.77 | 0.48 | -182.04 | 32.54 | 42.73 |
| 2 | $\beta_0, \beta_{CR}, \beta_{EV}, \beta_{RPE}, \gamma_{CR}, \gamma_{EV}, \gamma_{RPE}$ | 7 | 140.00 | 0.54 | -69.40 | 83.20 | 126.20 |
| 3 | $\beta_0, \beta_{CR}, \beta_{GR}, \gamma$ | 4 | 0 | 0.42 | 0 | 0 | 0 |
| 4 | $\beta_0, \beta_{CR}, \beta_{GR}, \gamma_{CR}, \gamma_{GR}$ | 5 | -146.81 | 0.48 | -272.15 | 26.37 | 98.97 |
| 5 | $\beta_0, \beta_{CR}, \gamma$ | 3 | 2395.62 | 0.18 | 1379.96 | 264.87 | 749.79 |
| 6 | $\beta_0, \beta_{GR}, \gamma$ | 3 | 403.46 | 0.34 | 228.69 | 21.24 | 153.52 |

Abbreviations: $\Delta$ BIC, Bayesian information criterion relative to the winning model in S<sup>+</sup> group (mM3); HC, healthy control; S<sup>-</sup>, patients without suicidal thoughts and behavior; S<sup>+</sup>, patients with suicidal thoughts and behavior.

In addition to the winning model (mM3) from S^+^ group, we also checked results from the classic mood model (mM1). Overall, we replicated previous findings (Figure S8): 1) mood sensitivity to CR, EV, and RPE were all significantly higher than 0 (*ps* < 0.001); 2) higher weight for RPE than EV (t = 5.760, *p* < 0.001). Although no significant group difference between S^+^ and S^-^ was found in each parameter (*ps* > 0.115), we replicated significant correlation between BSI-C and β_CR_ (rho = -0.239, *p* = 0.030). To explore why the classic mood model (mM1) did not outperform the CR-GR model, we examined expectation effect on mood, as previous literature showed impaired value expectation in patients with STB(60). Our data suggests a lower mood sensitivity to RPE relative to EV in S^+^ than HC (Figure S9; significant interaction between group and EV/RPE: F = 3.422, *p* = 0.035; with stronger mood sensitivity to RPE than EV in HC (F = 36.658, *p* < 0.001), while no such significant difference in S^+^ (F = 1.161, *p* = 0.283) and S^-^ (F = 3.009, *p* = 0.084)). Equal weights on EV and RPE suggest that expectations cancel out as RPE is the difference between the outcome and EV, resulting in outcome only. Then, we additionally fit a mood model with CR, GR, and EV components (Figure S9). We expect less negative mood sensitivity to EV in S^+^ than HC. As expected, in addition to replication of our main results (*ps* < 0.045; R^2^ for this model: 0.487), we observed a less negative mood sensitivity to EV in S^+^ than HC (t = 2.302, *p* = 0.023), which explains why the winning model shifts to mM3. Given that mM7 (splitting GR into better and worse terms) performed better than our winning model (mM3) in the S^-^ and HC groups, we also checked results from this model (Figure S10 and Table S6). Again, we found that S^+^ had significant lower β_CR_ than S^-^ and HC (for group effect: F = 44.660, *p* = 0.011; S^+^ vs. S^-^: t = -2.659, *p* = 0.009; S^+^ vs. HC: t = -2.589, *p* = 0.010; S^-^ vs. HC: t = 1.059, *p* = 0.292) and significant correlation between BSI-C and β_CR_, (rho = -0.297, *p* = 0.006) among patients, suggesting the robustness of mood sensitivity to certain reward in suicidal people. The marginally significant group effect in approach parameter (p = 0.053) remain marginally significant after correction (p=0.068). In addition to this, all results of interest, including gambling chosen, approach parameter, and mood sensitivity to CR, remained significant with FDR correction (ps <=0.05; Supplementary Note 8).

### Suicidal thought and behavior effect on gambling was mediated by mood sensitivity to certain rewards

To examine the association between risk behavior and atypical mood dynamics in suicidal patients, we then tested the correlation between participants’ gambling rate and mood sensitivity to certain reward (β_CR)_ in S^+^. We found significant negative correlation between gambling rate and β_CR_ in S^+^ (rho = -0.274, *p* = 0.037; Figure 3C), suggesting the lower mood sensitivity to certain reward, the more gambling behavior suicidal patients made. We did not observe such a significant correlation in S^-^ (rho = 0.246, *p* = 0.237) and there was significant correlational difference between S^+^ and S^-^ (Z = -2.109, *p* = 0.017; 42)), suggesting the suicidal-specific association of mood and choice.

### Replication of suicidal-related results in an independent dataset (n = 747)

Next, we collected online data on general volunteers to replicate our findings. In this large online dataset, we found lower mood experience in general volunteers who replied non-negatively to the Suicidal item of the BDI (S^+^). Regarding the initial mood rating (before the task), S^+^ exhibited significantly lower mood than S^-^ (t = -6.077, *p* < 0.001; Figure 4D). There was a trend for lower mood experience across time in S^+^ than S^-^ (t = - 1.600, *p* = 0.055; Figure 4E). Critically, we identified a significantly increased gambling behavior in S^+^ than S^-^, especially in the gain domain (t = 1.668, *p* = 0.048; Figure 4F). Approach-avoidance prospect theory model (mean pseudoR^2^ = 0.479) revealed a significantly heightened approach parameter in in S^+^ than S^-^ (t = 1.762, *p* = 0.039; Figure 4B), but not any other choice parameters (*ps* > 0.172). We also replicated the previous mediation result that STB increase risk behavior through stronger approach motivation (a ×b = 0.143, 95% CI = [0.016, 0.288], *p* = 0.031; Figure 4C). Regarding CR-GR mood model (mean R^2^ = 0.588), we observed significantly lower β_0_ in S^+^ than S^-^ (t = -2.018, *p* = 0.022; Figure 4F). Mood sensitivity to CR (t = -2.237, *p* = 0.013; Figure 4G), but not GR (t = -0. 187, *p* = 0.473; Figure 4G), was significantly reduced in S^+^ than S^-^. After controlling for depression severity using our established bifactor model (see ref 60 for details), these results remained significant (*ps* ≤ 0.050), except a marginally significant effect of group on gambling behavior (*p* = 0.059). Despite a trend, this effect with covariates of depression-related questionnaires is strong in our clinical cohort (*p* = 0.024). This suggests that the link between suicidality and risky behavior persists above and beyond general depressive symptoms.

**Figure 4.**
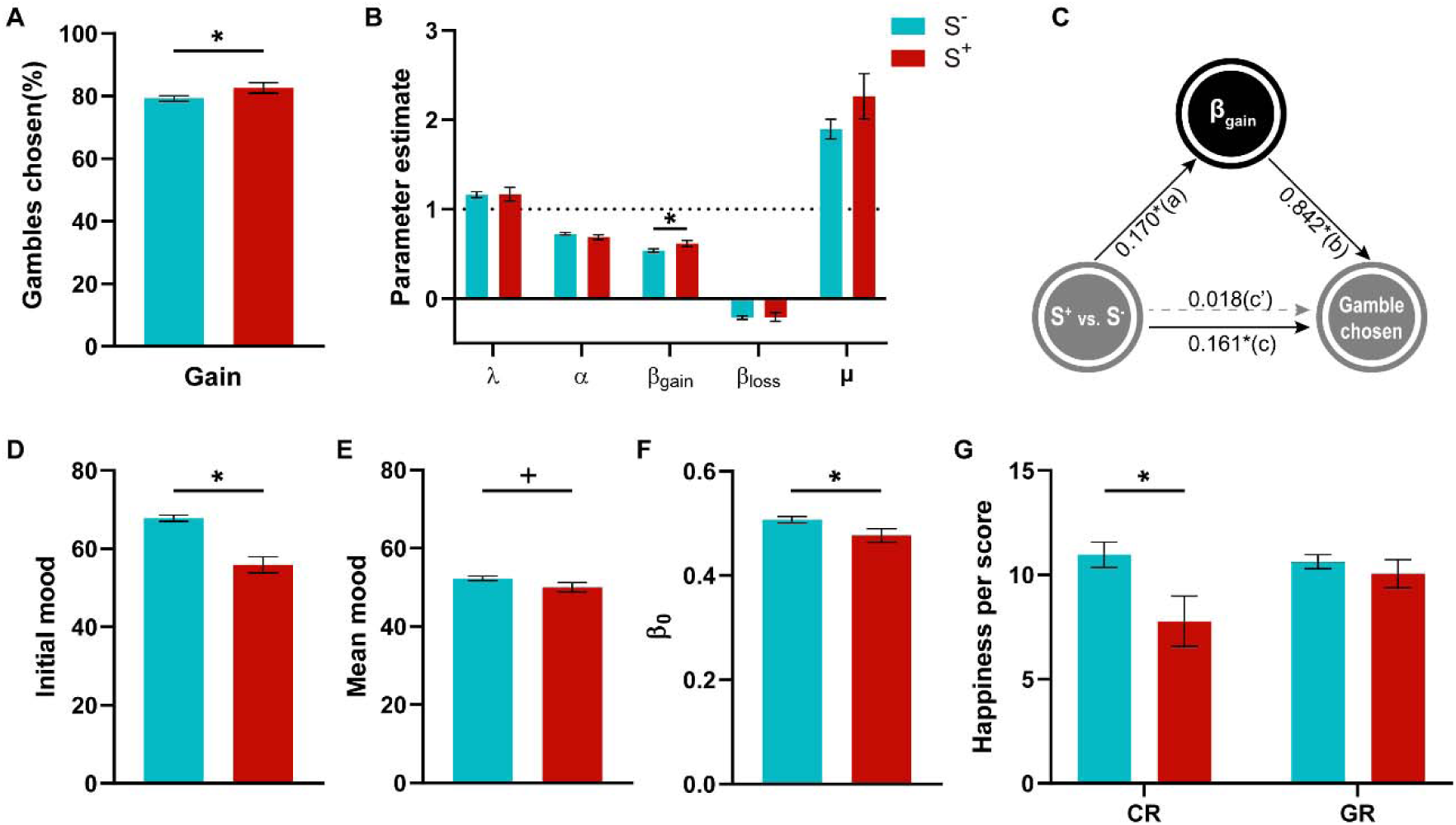
Validation of suicidal-related results in an independent dataset of general populations (n = 747). A) Group difference in gambling behavior in the gain domain. B) The estimated parameters from the winning choice model (pseudo R^2^ = 0.479) differed across groups, with higher approach behavior for S^+^. C) The mediation model among the group, *β_gain_*, and gambling behavior in the gain condition. The approach parameter mediated the group effect on increased gambling behavior in the gain condition. **D)** Group difference in mood before the task shows weakened mood in S^+^. E) Group difference in average mood displays lower mood experience in S^+^. FG) The estimated parameters from CR-GR mood model (mean R^2^ = 0.588). F) Group difference in mood baseline, β _0_. G) Group differences in mood sensitivity to certain reward (CR) and gamble reward (GR). Abbreviations: S^-^, general participants without suicidal thoughts and behavior; S^+^, general participants with suicidal thoughts and behavior; \**p*<0.05, ^+^ *p*<0.1.

These validation results suggest that our computational markers can generalize to general population. However, we did not observe any significant correlation between mood sensitivity to CR and gambling behavior (*ps* > 0.389), which suggests that the link between mood sensitivity to CR and gambling behavior may be specifically observable in suicidal patients. Alternatively, this non-replicated result may also reflect sample-specific or unstable effects, which needs to be interpreted with caution.

### Computational parameters were predictive of suicidal risks

To examine whether task-derived computational measures carry predictive information related to suicidal ideation (BSI-C) beyond single-parameter associations, we performed an additional multivariate prediction analysis using lasso regression within a cross-validation framework. Across 100 repetitions of 5-fold cross-validation, STB was significantly predicted by the computational parameters (mean *r* = 0.205, all *p_s_* < 0.039; Table 4), including approach motivation and mood sensitivity to certain rewards, across all participants, including both patients and healthy controls. Importantly, this predictive model generalized to the online sample (n = 747), where model-predicted scores were significantly correlated with scores on item 9 of BDI (r = 0.073, *p* = 0.045). We further confirmed the robustness of this predictive effect using 10-fold cross-validation, which yielded a similar pattern of results (Table 4). By contrast, predictive models based on choice-only or mood-only parameters did not show reliable generalization to the external validation sample (*ps* > 0.191), suggesting that the predictive signal emerged from the joint contribution of choice- and mood-related computational measures rather than either domain alone. In sum, these internal and external validation analyses corroborated the robustness of the predictive model of suicidal risks using computational markers.

**Table 4.**
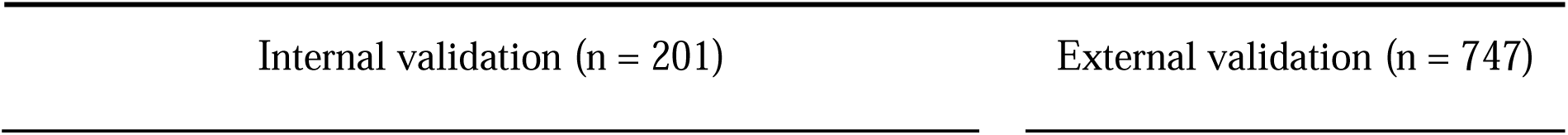
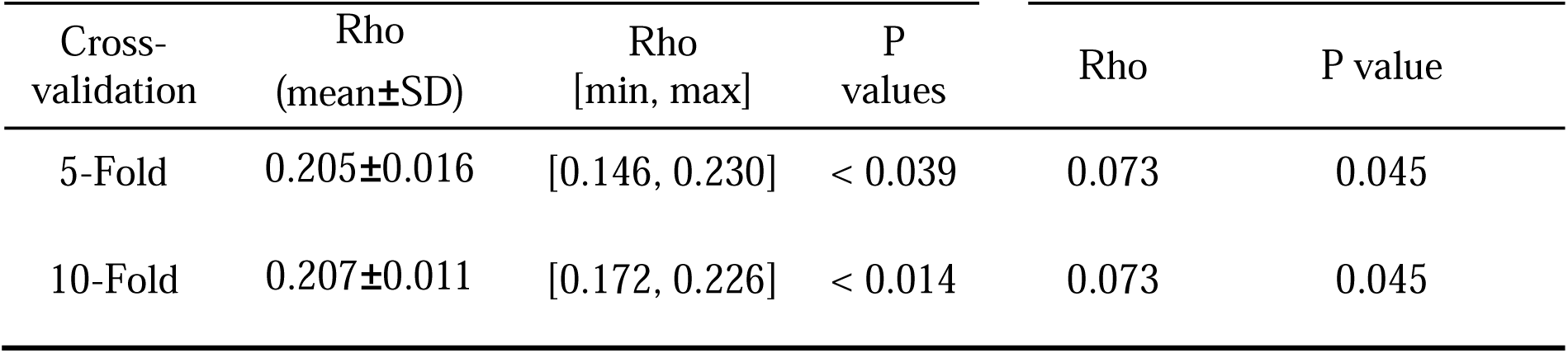
Suicidal risk prediction from computational parameters.

## Discussion

The current study tested cognitive and affective computational mechanisms for increased risk behavior in adolescent patients with suicidal thoughts and behaviors (STB), with a control group including adolescent patients without STB and sex/age-matched healthy control (HC). Firstly, we observed an increased gambling behavior and a lower overall mood in STB patients (S^+^), as compared to non-STB patients (S^-^) and HC, replicating previous findings(11–13,63). Secondly, using an approach-avoidance prospect theory model, we found heightened approach motivation in S^+^ than S^-^ and HC, which explained increased gambling choices for STB, suggesting an over-reactivity of the approach system to approach risky options. Thirdly, using a momentary mood model, we showed that lower mood sensitivity to certain outcomes in S^+^ compared to S^-^ and HC, which was driven by lower mood sensitivity to certain outcomes in S^+^ than S^-^ and HC. These computational markers generalized to general population (*n*□=□747). Importantly, mood hyposensitivity to certain reward specifically correlated to more gambling behavior in S^+^, offering a mood computational account for increased risk behavior in STB. Beyond these specific findings, this work highlights the broader utility of combining computational modelling with momentary mood measures to better characterize behavioral differences relevant to psychiatric symptoms, and to show how choice and mood data can jointly inform our understanding of psychiatric phenomena.

Our results suggest a unique reason for the twofold observations that STB patients display an increase in both risk taking and impulsivity, defined as a tendency to act quickly without planning while failing to inhibit a behavior that is likely to result in negative consequences(64–68). Indeed, we did not observe a difference in risk attitude (e.g., risk aversion and loss aversion) per se between STB and controls but instead a higher approach behavior towards largest rewards (i.e., the lotteries) in STB patients. This would result from the value-independent term in the model that represents forms of approach in the face of gains(17,69,70). Such approach actions are elicited without regard to their actual contingent benefits and therefore correspond to impulsive behavior. A substantial body of research has shown that impulsivity, as assessed either through questionnaires or clinical observations, is a key predictor for STB (for a review, see Franklin et.al (2016)). Our study employed computational modeling to quantitively elucidate the altered approach-system processing for increased risky behavior in STB, offering enhanced predictive power and generalizability (39). On the other hand, contrary to the proposal of atypical avoidance system(20,23), we did not observe significant group difference in avoidance, which may be attributed to the different involvement of the motivational system in learning and non-leaning contexts(18,71). In our model specification, motivational systems work in a value independent way in the non-learning context. Consistent with the view that suicide is an escape from intolerable affective states(3), risky behavior in suicidal individuals may be rewarding. In clinical practices, understanding the distortion of the approach system in STB may encourage mental health professionals to closely monitor patients who exhibit heightened approach tendencies.

Such vigilance may enable early detection of risk-related behaviors, thus facilitating timely intervention strategies tailored to mitigate impulsivity-driven actions that may elevate the likelihood of STB.

Consistent with suicidal-related theories(3,72,73) and as summarized by Millner et.al (2020), we observed lower mood levels in patients with STB, regarding both initial happiness and mood baseline (the latter corresponding to the steady state mood converges to). More importantly, STB patients’ mood was less sensitive to certain outcomes than control without STB, which would lead them to take more risk regardless of the gain at stake and therefore to potentially experience more suboptimal outcomes than controls(11). Although no direct causal link was established between STB and happiness ratings in response to wins or losses, recent literature has documented associations between STB and anhedonia symptoms (albeit with mixed evidence; for a review, see (73)), where anhedonia can be assessed through affective reactivity to wins versus losses ((74)). Our findings thus provide support for the presence of anhedonia in STB, particularly in response to certain outcomes. Surprisingly, mood model-based analysis did not support the effect of expectations and prediction errors on mood in healthy people (the “CR-EV-RPE model”(17,28,75)), but suggest instead a dissociation between certain outcomes and lottery outcomes (the “CR-GR model”). These two models differed with respect to the inclusion of reward expectation terms, the former including it unlike the latter. This difference can be explained by the lower expected value signal in patients with STB(60), resulting in insufficient expectation representations of the gamble option to influence mood dynamics. An alternative explanation could be the duration of the chosen option display which was considerably lower in our design than in other mood studies (e.g., 0.5 s in our study versus 6 s in (28)), which would not leave enough time for expectation to be built. It is also possible that the current winning model was specific to adolescents. Given that Rutledge et al., (2017) supported the “CR-EV-RPE model” in adults with depression, our study with adolescent populations may suggest a developmental change for mood sensitivities. Within the winning CR-GR model, we observed that S^+^ specifically exhibited lower mood sensitivity to CR than GR, which was driven by mood hyposensitivity to CR in S^+^ than S^-^ and HC. This mood insensitivity was associated with STB severity, which was replicated when using the CR-EV-RPE model. Importantly, we found that mood hyposensitivity to certain reward was specifically correlated to gambling behavior in patients with STB, suggesting the potential mood computational mechanism for increased risk behavior in STB. As for clinical practices, CR-based anhedonia linked to CR (computational reactivity) in STB may prompt mental health professionals to closely monitor patients who exhibit mood insensitivity to certain daily events. This proactive monitoring could aid in identifying and addressing risk-related behaviors early on.

With replication in an independent dataset with large sample size (n = 747), this study provides robust evidence of the affective and cognitive computational mechanisms underlying heightened risky behavior in adolescents with STB. In addition, these results remained significant after controlling for demographics, social and clinical variables, medication factors, and the timing of suicidal events (Supplementary Note 3 & 4). However, this study could not differentiate between suicidal thoughts and suicidal behaviors. Although it has been shown that they represented different decision-making processes with different neural underpinnings (76–78), our data did not reveal significant differences between them (see Supplementary Note 2). Future research would benefit from examining these distinctions at the neural level. Nonetheless, by combining the suicidal ideation and suicidal attempt groups into a single STB group (40–45), our findings highlight why adolescents with suicidality exhibit a preference for risky behavior. These findings carry important clinical implications for early prevention of adolescent suicidality. Notably, this study, like many traditional studies on suicidality (40–42,79,80), does not seek to elucidate the affective and cognitive mechanisms underlying fluctuations in suicidal thoughts. Given the inherently variable nature of suicidal ideation, recent research has increasingly adopted ecological momentary assessments to capture real-time variations in suicidal ideations(45,81,82). While such methods can help predict when suicidal ideation may arise, they fall short of explaining the underlying mechanisms driving these thoughts. In contrast, our approach, consistent with traditional literature (40–44,79), is directed at understanding why individuals with STB are more inclined toward risky behavior. We acknowledge the interaction between environmental stressor and the occurrence of STB, noting that suicidal severity often diminishes once the stressor is removed (44,83). This is a crucially important issue in current psychiatric research. For instance, patients with MDD sometimes experience depressive episodes, particularly in response to stressful events. However, collecting data during STB is both impractical and ethically challenging. Our grouping approach assumes of trait-driven STB: individuals with a history of STB, despite not during the experiment, represent a cluster of suicidal-related traits (83,84). Sensitivity analyses for STB timeframe support this assumption (see Supplementary Note 3). We also recognize that these affective and cognitive impairments may worsen under stress(44,83). Future studies would benefit from investigating how acute stress influences the propensity for risky behavior in individuals with STB.

Given that STB is a challenging multifactorial phenomenon, the development of a formal theory to quantify suicide seems necessary(20,21,85). Our cognitive and affective computational insights may pave the way for such a formal theory. Although previous literature has shown various cognitive impairments(13), e.g., executive function, in STB(86), our work is the first to quantify mood dynamics impairment and their behavioral consequences, providing insight into potential target to prevent and intervene STB. Our results indeed provide a computational mechanism for the main theories of suicide, linking low mood to suicidal behaviors. Suicide behavior is conceived to result from an intention shaped by various motivational factors (e.g., feeling of entrapment, belongness, burdensomeness(87)). The suicidal intent may then progress to suicidal behavior, which is thought to be moderated by impulsive decisions (e.g., (88)). A possibility is that the approach component becomes excessive as the suicidal intent emerges. These findings provide new insights into the putative dynamics underpinning STB, and offer potential markers for the early prediction, screening, detection, and prevention of suicidal behavior. These results would explain the observed increase in risk-taking behavior in STB such as substance use, early onset of sexual intercourse and physical fighting independent of psychiatric diagnosis.

Several limitations are worth mentioning. First, our cross-section findings are of correlational nature. Causal relationships remain to be tested in a longitudinal study. Second, although we assumed that increased risky behavior in STB was suboptimal, the current task was not suited to test this, given the task design of random feedback for gambling option. Future work in learning paradigms, where optimality is well defined, may be better suited to test earnings-based links to STB. Third, despite replicating our main results in an independent dataset (n=747), the modest S^−^ subgroup size (n=25) has a limited statistical power. Next, we did not evaluate the noise in our estimate e.g., by assessing the test-retest reliability on the task parameters and it is indeed possible that parameter estimate is somehow noisy.

To conclude, this study examined cognitive and affective computational mechanisms underlying increased risk behavior in adolescent patients with suicidal thoughts and behaviors. Given very limited predictive abilities of suicide from previous risk-factor investigations(8), our study offers a potential new perspective of mood, at the core of STB, and reveals a relationship between low mood sensitivity to certain reward and an increased risk behavior in STB and possibly suggesting dysfunctional dopaminergic and serotoninergic systems. Our findings suggest that computational measures may capture variance related to suicidal tendency in adolescents and thus may be relevant for future work on early identification and prevention of suicidality.

## Data Availability

All data produced in the present study are available upon reasonable request to the authors

## Acknowledgements

This study was funded by the National Natural Science Foundation of China (31920103009,62173069,62006038, 32500929), the Major Project of National Social Science Foundation (20&ZD153), Ministry of Education Humanities and Social Sciences (25YJC190023), Shenzhen-Hong Kong Institute of Brain Science – Shenzhen Fundamental Research Institutions (2019SHIBS0003),Science and Technology Bureau of Chengdu Program (2022-YF09-00023-SN),Sichuan Province Science and Technology Support Program (2022YFS0180), and Guangdong Provincial Advanced Education Institutions Young Innovative Talent Project (2025WQNCX013).

## Data availability

The data that support the findings of this study are available from the corresponding author upon reasonable request.

## References

1. Preventing suicide: A global imperative (2014): World Health Organization.

2. Hawton K, Saunders KE, O’Connor RC (2012): Self-harm and suicide in adolescents. The Lancet 379: 2373–2382.

3. O’Connor RC, Nock MK (2014): The psychology of suicidal behaviour. Lancet Psychiatry 1: 73–85.

4. Guze SB, Robins E (1970): Suicide and Primary Affective Disorders. British Journal of Psychiatry 117: 437–438.

5. Franklin JC, Ribeiro JD, Fox KR, Bentley KH, Kleiman EM, Huang X, et al. (2017): Risk factors for suicidal thoughts and behaviors: A meta-analysis of 50 years of research. Psychol Bull 143: 187–232.

6. King CA, Arango A, Ewell Foster C (2018): Emerging trends in adolescent suicide prevention research. Curr Opin Psychol 22: 89–94.

7. Schmaal L, van Harmelen A-L, Chatzi V, Lippard ETC, Toenders YJ, Averill LA, et al. (2020): Imaging suicidal thoughts and behaviors: a comprehensive review of 2 decades of neuroimaging studies. Mol Psychiatry 25: 408–427.

8. Franklin JC, Ribeiro JD, Fox KR, Bentley KH, Kleiman EM, Huang X, et al. (2017): Risk factors for suicidal thoughts and behaviors: A meta-analysis of 50 years of research. Psychol Bull 143: 187–232.

9. da Silva AG, Malloy-Diniz LF, Garcia MS, Figueiredo CGS, Figueiredo RN, Diaz AP, Palha AP (2018): Cognition As a Therapeutic Target in the Suicidal Patient Approach. Front Psychiatry 9. 10.3389/fpsyt.2018.00031

10. Oldershaw A, Simic M, Grima E, Jollant F, Richards C, Taylor L, Schmidt U (2012): The Effect of Cognitive Behavior Therapy on Decision Making in Adolescents who Self-Harm: A Pilot Study. Suicide Life Threat Behav 42: 255–265.

11. Sastre-Buades A, Alacreu-Crespo A, Courtet P, Baca-Garcia E, Barrigon ML (2021): Decision-making in suicidal behavior: A systematic review and meta-analysis. Neurosci Biobehav Rev 131: 642–662.

12. Perrain R, Dardennes R, Jollant F (2021): Risky decision-making in suicide attempters, and the choice of a violent suicidal means: an updated meta-analysis. J Affect Disord 280: 241–249.

13. Richard-Devantoy S, Berlim MT, Jollant F (2014): A meta-analysis of neuropsychological markers of vulnerability to suicidal behavior in mood disorders. Psychol Med 44: 1663–1673.

14. Liu Q, Zhong R, Ji X, Law S, Xiao F, Wei Y, et al. (2022): Decision-making biases in suicide attempters with major depressive disorder: A computational modeling study using the balloon analog risk task (BART). Depress Anxiety 39: 845–857.

15. Baek K, Kwon J, Chae J-H, Chung YA, Kralik JD, Min J-A, et al. (2017): Heightened aversion to risk and loss in depressed patients with a suicide attempt history. Sci Rep 7: 11228.

16. Alacreu-Crespo A, Guillaume S, Sénèque M, Olié E, Courtet P (2020): Cognitive modelling to assess decision-making impairments in patients with current depression and with/without suicide history. European Neuropsychopharmacology 36: 50–59.

17. Rutledge RB, Skandali N, Dayan P, Dolan RJ (2015): Dopaminergic Modulation of Decision Making and Subjective Well-Being. Journal of Neuroscience 35: 9811–9822.

18. Rutledge RB, Smittenaar P, Zeidman P, Brown HR, Adams RA, Lindenberger U, et al. (2016): Risk Taking for Potential Reward Decreases across the Lifespan. Current Biology 26: 1634–1639.

19. Corr PJ, McNaughton N (2012): Neuroscience and approach/avoidance personality traits: A two stage (valuation–motivation) approach. Neurosci Biobehav Rev 36: 2339–2354.

20. Dombrovski AY, Hallquist MN (2022): Search for solutions, learning, simulation, and choice processes in suicidal behavior. WIREs Cognitive Science 13. 10.1002/wcs.1561

21. Karvelis P, Diaconescu AO (2022): A Computational Model of Hopelessness and Active-Escape Bias in Suicidality. Computational Psychiatry 6: 34.

22. Millner AJ, den Ouden HEM, Gershman SJ, Glenn CR, Kearns JC, Bornstein AM, et al. (2019): Suicidal thoughts and behaviors are associated with an increased decision-making bias for active responses to escape aversive states. J Abnorm Psychol 128: 106–118.

23. Dombrovski AY, Hallquist MN (2017): The decision neuroscience perspective on suicidal behavior. Curr Opin Psychiatry 30: 7–14.

24. Van Orden KA, Witte TK, Cukrowicz KC, Braithwaite SR, Selby EA, Joiner TE (2010): The interpersonal theory of suicide. Psychol Rev 117: 575–600.

25. O’Connor RC, Cleare S, Eschle S, Wetherall K, Kirtley OJ (2016): The Integrated Motivational-Volitional Model of Suicidal Behavior. The International Handbook of Suicide Prevention. Chichester, UK: John Wiley & Sons, Ltd, pp 220–240.

26. Klonsky ED, May AM (2015): The Three-Step Theory (3ST): A New Theory of Suicide Rooted in the “Ideation-to-Action” Framework. Int J Cogn Ther 8: 114–129.

27. Millner AJ, Robinaugh DJ, Nock MK (2020): Advancing the Understanding of Suicide: The Need for Formal Theory and Rigorous Descriptive Research. Trends Cogn Sci 24: 704–716.

28. Rutledge RB, Skandali N, Dayan P, Dolan RJ (2014): A computational and neural model of momentary subjective well-being. Proceedings of the National Academy of Sciences 111: 12252–12257.

29. Kao C-H, Feng GW, Hur JK, Jarvis H, Rutledge RB (2023): Computational models of subjective feelings in psychiatry. Neurosci Biobehav Rev 145: 105008.

30. Emanuel A, Eldar E (2023): Emotions as computations. Neurosci Biobehav Rev 144: 104977.

31. Eldar E, Rutledge RB, Dolan RJ, Niv Y (2016): Mood as representation of momentum. Trends Cogn Sci 20: 15–24.

32. Schiller D, Yu ANC, Alia-Klein N, Becker S, Cromwell HC, Dolcos F, et al. (2024): The Human Affectome. Neurosci Biobehav Rev 158: 105450.

33. Rutledge RB, de Berker AO, Espenhahn S, Dayan P, Dolan RJ (2016): The social contingency of momentary subjective well-being. Nat Commun 7: 11825.

34. Blain B, Rutledge RB (2020): Momentary subjective well-being depends on learning and not reward. Elife 9. 10.7554/eLife.57977

35. Rutledge RB, Skandali N, Dayan P, Dolan RJ (2014): A computational and neural model of momentary subjective well-being. Proceedings of the National Academy of Sciences 111: 12252–12257.

36. Rutledge RB, Moutoussis M, Smittenaar P, Zeidman P, Taylor T, Hrynkiewicz L, et al. (2017): Association of Neural and Emotional Impacts of Reward Prediction Errors With Major Depression. JAMA Psychiatry 74: 790.

37. Csukly G, Farkas K, Fodor T, Unoka Z, Polner B (2023): Stronger coupling of emotional instability with reward processing in borderline personality disorder is predicted by schema modes. Psychol Med 53: 6714–6723.

38. Wang Z, Nan T, Goerlich KS, Li Y, Aleman A, Luo Y, Xu P (2023): Neurocomputational mechanisms underlying fear-biased adaptation learning in changing environments. PLoS Biol 21: e3001724.

39. Huys QJM, Maia T V, Frank MJ (2016): Computational psychiatry as a bridge from neuroscience to clinical applications. Nat Neurosci 19: 404–413.

40. Glenn CR, Millner AJ, Esposito EC, Porter AC, Nock MK (2019): Implicit Identification with Death Predicts Suicidal Thoughts and Behaviors in Adolescents. Journal of Clinical Child & Adolescent Psychology 48: 263–272.

41. Glenn JJ, Werntz AJ, Slama SJK, Steinman SA, Teachman BA, Nock MK (2017): Suicide and self-injury-related implicit cognition: A large-scale examination and replication. J Abnorm Psychol 126: 199–211.

42. Millner AJ, den Ouden HEM, Gershman SJ, Glenn CR, Kearns JC, Bornstein AM, et al. (2019): Suicidal thoughts and behaviors are associated with an increased decision-making bias for active responses to escape aversive states. J Abnorm Psychol 128: 106–118.

43. Miller AB, Jenness JL, Elton AL, Pelletier-Baldelli A, Patel K, Bonar A, et al. (2024): Neural Markers of Emotion Reactivity and Regulation Before and After a Targeted Social Rejection: Differences Among Girls With and Without Suicidal Ideation and Behavior Histories. Biol Psychiatry 95: 1100–1109.

44. Eisenlohr-Moul TA, Miller AB, Giletta M, Hastings PD, Rudolph KD, Nock MK, Prinstein MJ (2018): HPA axis response and psychosocial stress as interactive predictors of suicidal ideation and behavior in adolescent females: a multilevel diathesis-stress framework. Neuropsychopharmacology 43: 2564–2571.

45. Miller AB, Eisenlohr-Moul T, Giletta M, Hastings PD, Rudolph KD, Nock MK, Prinstein MJ (2017): A within-person approach to risk for suicidal ideation and suicidal behavior: Examining the roles of depression, stress, and abuse exposure. J Consult Clin Psychol 85: 712–722.

46. Wang Z, Wang T, Nan T, Xu J, Aleman A, Luo Y, et al. (2025, June 23): Dissociable Roles of Reward Prediction Error in the Contrasting Mood Dynamics of Depression and Anxiety. 10.31234/osf.io/z6s2r_v1

47. Zhang J, Brown GK (2007): Psychometric Properties of the Scale for Suicide Ideation in China. Archives of Suicide Research 11: 203–210.

48. Zhao X, Zhang Y, Li L, Zhou Y (2005): Evaluation on reliability and validity of Chinese version of childhood trauma questionnaire. Chinese Journal of Tissue Engineering Research 209–211.

49. Zhu X, Auerbach RP, Yao S, Abela JRZ, Xiao J, Tong X (2008): Psychometric properties of the Cognitive Emotion Regulation Questionnaire: Chinese version. Cogn Emot 22: 288–307.

50. Shek DTL (1988): Reliability and factorial structure of the Chinese version of the State-Trait Anxiety Inventory. J Psychopathol Behav Assess 10: 303–317.

51. Shek DTL (1990): Reliability and factorial structure of the chinese version of the Beck Depression Inventory. J Clin Psychol 46: 35–43.

52. Jiang L, Wang Y, Zhang Y, Li R, Wu H, Li C, et al. (2019): The Reliability and Validity of the Center for Epidemiologic Studies Depression Scale (CES-D) for Chinese University Students. Front Psychiatry 10. 10.3389/fpsyt.2019.00315

53. Kahneman, L. DANIE, Tversky A (1979): Prospect theory: An analysis of decision under risk. Econometrica 47: 363–391.

54. Sokol-Hessner P, Rutledge RB (2019): The Psychological and Neural Basis of Loss Aversion. Curr Dir Psychol Sci 28: 20–27.

55. Rutledge RB, Skandali N, Dayan P, Dolan RJ (2015): Dopaminergic modulation of decision making and subjective well-being. Journal of Neuroscience 35: 9811–9822.

56. Wang Z, Nan T, Goerlich KS, Li Y, Aleman A, Luo Y, Xu P (2023): Neurocomputational mechanisms underlying fear-biased adaptation learning in changing environments ((M. F. S. Rushworth, editor)). PLoS Biol 21: e3001724.

57. Blain B, Rutledge RB (2020): Momentary subjective well-being depends on learning and not reward. Elife 9. 10.7554/eLife.57977

58. Jangraw DC, Keren H, Sun H, Bedder RL, Rutledge RB, Pereira F, et al. (2023): A highly replicable decline in mood during rest and simple tasks. Nat Hum Behav 7: 596–610.

59. Rutledge RB, Moutoussis M, Smittenaar P, Zeidman P, Taylor T, Hrynkiewicz L, et al. (2017): Association of Neural and Emotional Impacts of Reward Prediction Errors With Major Depression. JAMA Psychiatry 74: 790.

60. Dombrovski AY, Szanto K, Clark L, Reynolds CF, Siegle GJ (2013): Reward Signals, Attempted Suicide, and Impulsivity in Late-Life Depression. JAMA Psychiatry 70: 1020.

61. Lenhard W, Lenhard A (2014): Hypothesis Tests for Comparing Correlations. Https://Www.Psychometrica.de/Correlation.Html.

62. Jollant F, Bellivier F, Leboyer M, Astruc B, Torres S, Verdier R, et al. (2005): Impaired Decision Making in Suicide Attempters. American Journal of Psychiatry 162: 304–310.

63. Ortin A, Lake AM, Kleinman M, Gould MS (2012): Sensation seeking as risk factor for suicidal ideation and suicide attempts in adolescence. J Affect Disord 143: 214–222.

64. Horesh N, Gothelf D, Ofek H, Weizman T, Apter A (1999): Impulsivity as a correlate of suicidal behavior in adolescent psychiatric inpatients. Crisis: The Journal of Crisis Intervention and Suicide Prevention 20: 8–14.

65. Javdani S, Sadeh N, Verona E (2011): Suicidality as a function of impulsivity, callous–unemotional traits, and depressive symptoms in youth. J Abnorm Psychol 120: 400–413.

66. Kingsbury S, Hawton K, Steinhardt K, James A (1999): Do Adolescents Who Take Overdoses Have Specific Psychological Characteristics? A Comparative Study With Psychiatric and Community Controls. J Am Acad Child Adolesc Psychiatry 38: 1125–1131.

67. Renaud J, Berlim MT, McGirr A, Tousignant M, Turecki G (2008): Current psychiatric morbidity, aggression/impulsivity, and personality dimensions in child and adolescent suicide: A case-control study. J Affect Disord 105: 221–228.

68. Dayan P, Niv Y, Seymour B, D. Daw N (2006): The misbehavior of value and the discipline of the will. Neural Networks 19: 1153–1160.

69. Bushong B, King LM, Camerer CF, Rangel A (2010): Pavlovian Processes in Consumer Choice: The Physical Presence of a Good Increases Willingness-to-Pay. American Economic Review 100: 1556–1571.

70. Guitart-Masip M, Huys QJM, Fuentemilla L, Dayan P, Duzel E, Dolan RJ (2012): Go and no-go learning in reward and punishment: Interactions between affect and effect. Neuroimage 62: 154–166.

71. Allen NB, Nelson BW, Brent D, Auerbach RP (2019): Short-term prediction of suicidal thoughts and behaviors in adolescents: Can recent developments in technology and computational science provide a breakthrough? J Affect Disord 250: 163–169.

72. Stewart JG, Polanco-Roman L, Duarte CS, Auerbach RP (2019): Neurocognitive Processes Implicated in Adolescent Suicidal Thoughts and Behaviors: Applying an RDoC Framework for Conceptualizing Risk. Curr Behav Neurosci Rep 6: 188–196.

73. Hall AF, Browning M, Huys QJM (2024): The computational structure of consummatory anhedonia. Trends Cogn Sci 28: 541–553.

74. Vanhasbroeck N, Devos L, Pessers S, Kuppens P, Vanpaemel W, Moors A, Tuerlinckx F (2021): Testing a computational model of subjective well-being: a preregistered replication of Rutledge et al. (2014). Cogn Emot 35: 822–835.

75. Wagner G, Li M, Sacchet MD, Richard-Devantoy S, Turecki G, Bär K-J, et al. (2021): Functional network alterations differently associated with suicidal ideas and acts in depressed patients: an indirect support to the transition model. Transl Psychiatry 11: 100.

76. Schmaal L, van Harmelen A-L, Chatzi V, Lippard ETC, Toenders YJ, Averill LA, et al. (2020): Imaging suicidal thoughts and behaviors: a comprehensive review of 2 decades of neuroimaging studies. Mol Psychiatry 25: 408–427.

77. Saffer BY, Klonsky ED (2018): Do neurocognitive abilities distinguish suicide attempters from suicide ideators? A systematic review of an emerging research area. Clinical Psychology: Science and Practice 25: e12227.

78. Jollant F, Colle R, Nguyen TML, Corruble E, Gardier AM, Walter M, et al. (2023): Ketamine and esketamine in suicidal thoughts and behaviors: a systematic review. Ther Adv Psychopharmacol 13. 10.1177/20451253231151327

79. Tsypes A, Hallquist MN, Ianni A, Kaurin A, Wright AGC, Dombrovski AY (2024): Exploration-Exploitation and Suicidal Behavior in Borderline Personality Disorder and Depression. JAMA Psychiatry 81: 1010.

80. Kleiman EM, Turner BJ, Fedor S, Beale EE, Huffman JC, Nock MK (2017): Examination of real-time fluctuations in suicidal ideation and its risk factors: Results from two ecological momentary assessment studies. J Abnorm Psychol 126: 726–738.

81. Wang S, Van Genugten RDI, Yacoby Y, Pan W, Bentley KH, Bird SA, et al. (2024): Building personalized machine learning models using real-time monitoring data to predict idiographic suicidal thoughts. Nature Mental Health. 10.1038/s44220-024-00335-w

82. Stewart JG, Shields GS, Esposito EC, Cosby EA, Allen NB, Slavich GM, Auerbach RP (2019): Life Stress and Suicide in Adolescents. J Abnorm Child Psychol 47: 1707–1722.

83. Dombrovski AY, Hallquist MN (2022): Search for solutions, learning, simulation, and choice processes in suicidal behavior. WIREs Cognitive Science 13. 10.1002/wcs.1561

84. Millner AJ, Robinaugh DJ, Nock MK (2020): Advancing the Understanding of Suicide: The Need for Formal Theory and Rigorous Descriptive Research. Trends Cogn Sci 24: 704–716.

85. Bredemeier K, Miller IW (2015): Executive function and suicidality: A systematic qualitative review. Clin Psychol Rev 40: 170–183.

86. Klonsky ED, Saffer BY, Bryan CJ (2018): Ideation-to-action theories of suicide: a conceptual and empirical update. Curr Opin Psychol 22: 38–43.

87. O’Connor RC, Kirtley OJ (2018): The integrated motivational–volitional model of suicidal behaviour. Philosophical Transactions of the Royal Society B: Biological Sciences 373: 20170268.

