## Supplement for "Mood computational mechanisms underlying increased risk behavior in adolescent suicidal patients"

**Supplementary Note 1: Sample characteristics**

There was a lack of consensual definition for suicidal thoughts and behaviors (STB; Jollant et al., 2023). See Goodfellow et al., (2018) for a systematic review for nomenclatures of suicidology. In the current study, the threshold for suicidal ideation was active thoughts of suicide, i.e., wishing to die and having some intention to do so, whilst suicide attempt was characterized by a deliberate action taken to end one’s life. Consistent with prior suicidal research (C. R. Glenn et al., 2019; Miller et al., 2024; Millner et al., 2019), STB referred to individuals either with suicidal ideation or suicidal attempt. STB has been understood as the transdiagnostic symptom. Indeed, literature reported that STB co-occurred with many mental disorders, such as major depressive disorders (MDD), generalized anxiety disorders (GAD), bipolar disorders (BD), schizophrenia (Pompili et al., 2007), borderline personality disorders (Tsypes et al., 2024), and substance use (Poorolajal et al., 2016), amongst others. Given that many intervention studies of STB mainly focused on mood and anxiety disorders (see (Jollant et al., 2023) for a review) and adolescence is an important time window for the emergence of affective problems (O’Connor & Nock, 2014), we thus recruited adolescent patients who were currently diagnosed with MDD, GAD, or BD. Patients were diagnosed by two experienced psychiatrists using the Structured Clinical Interview for DSM-IV-TR-Patient Edition (SCID-P, 2/2001 revision). It is important to note there was no adolescent version of SCID. Therefore, experienced psychiatrists made diagnostic decisions with appropriate adjustments for adolescent populations. Despite not conducting a structured interview to directly assess suicidal risk, the psychiatrists remained vigilant for signs of suicidal ideation and inquired about suicidal-related thoughts and behaviors from both the patients and their families. If the patient openly discussed suicide, the doctor asked follow-up questions, like “Have you thought about how you would harm yourself?” or “do you have a specific plan in mind?”. Instead, if the patient did not bring up suicide, the psychiatrist could gently probe with questions, such as “Have you been feeling overwhelmed or hopeless lately?” or “Do you ever feel like life is no longer worth living” to assess potential suicidal thoughts and behaviors.

**Supplementary Note 2: Suicidal Attempts vs. Suicidal ideations**

Consistent with previous suicidal-related literature ((Eisenlohr-Moul et al., 2018; C. R. Glenn et al., 2019; J. J. Glenn et al., 2017; Miller et al., 2017, 2024; Millner et al., 2019), this study focused on why adolescent patients with STB showed more risky behavior. By dividing patients into two groups: patient with STB (S^+^) and without STB (S^-^), this work revealed the general tendency for suicidal risks and had clinical implications for suicidal prevention, especially among adolescence. Although not at the heart of this study, as a secondary analysis, we checked differences in patients with suicidal attempts (SA) and without SA. To carefully control for suicidal ideations (SI), we mainly checked differences between SA and suicidal ideation (SI) groups. However, results showed no significant group difference in either choice or mood indices (*ps* > 0.148; Figure S1).

**Supplementary Note 3: Sensitivity analysis for suicidal timeframe**

In this study, we focused on adolescents (age: 10-19 years) who made suicidal thoughts and behaviors during their adolescent period (10-19 years). The longest timeframe for STB was 6 years (mean: 14.6 months; median: 6 months; standard deviation: 17.74). If we limited the timeframe, e.g., midpoint of the longest timeframe (3 years; 36 months), all results, including increased risky behavior, heightened approach motivation, and lower mood sensitivity to certain reward in STB, remained significant (*ps* < 0.026).

**Supplementary Note 4: Control analysis for childhood maltreatment, emotion regulation, and depression/anxiety symptoms**

It has been shown that patients with STB have childhood maltreatment problems, inability to regulate their emotions adaptively, and high levels of depression and anxiety per se (Neacsiu et al., 2018; Sarchiapone et al., 2007). Indeed, we observed these patterns in our data (see Table 1). To check whether our current effects were specific to suicidal thoughts and behaviors, but not these symptoms, we performed control analysis for these variables. Specifically, we used median split to check each potential confound on the gambling chosen, approach parameter, and mood sensitivity to certain reward. Results showed no significant effect in the variables of interests (*ps* > 0.056; Figure S3 & S4). In addition, our recent work identified the specific influence of depression/anxiety on mood sensitivity to reward prediction error, but not mood sensitivity to certain reward (Wang et al., 2025), further suggesting that the current group differences were specific to suicidal thoughts and behaviors, but not general severity of internalizing psychopathology.

**Supplementary Note 5: Justify for the modeling technique.**

**Model recovery**: We performed simulation-based model recovery analyses over 1,000 iterations. In each iteration, the task design matrix was randomly shuffled to vary trial order while maintaining the original task structure. For choice model recovery, for each of the three candidate choice models, a random parameter set was generated and used to simulate choices based on the shuffled design matrix. Each simulated dataset was then fit with all three choice models, and BIC was computed for each fit. The model with the lowest BIC was selected as the winning model. Recovery performance was summarized in a confusion matrix, where each cell reflected the proportion of times a given model was selected when data were generated from a particular model.

For mood model recovery, simulated outcomes were first generated from the fitted choice model and then used to simulate mood ratings. For each of the six candidate mood models, random parameters were generated and used to produce synthetic mood data. Mood model recovery was performed in a stage-by-stage manner, mirroring the model comparison procedure used in the main analyses because each model comparison step addresses a specific question and avoids dispersion of model evidence across multiple similar models within over-represented model families, which can reduce apparent recovery of the true generative model. Specifically, rather than entering all mood models into a single recovery space, recovery was evaluated separately within each comparison space used for model selection. Thus, each confusion matrix reflects recovery among only the models that were directly compared at that stage. Note that the winning model was determined at individual level. As shown in Figure S5, model identifiability was high overall, indicating good recovery performance for both the choice and mood models.

**Parameter recovery:** To assess parameter recovery, the generating parameters were further correlated with the recovered parameters from the corresponding fitted model. Note that simulated datasets were not generated from parameters estimated from empirical data. Instead, parameter values were randomly drawn from the predefined bounded parameter space used for model fitting. This approach was adopted to minimize the correlation structure inherited from the empirical fits and to promote greater orthogonality among parameters, thereby providing a cleaner test of parameter recoverability. Figure S6 shows good parameter recovery for both choice and mood winning model (choice: rs > 0.37, *ps* < 0.001; intraclass coefficients > 0.24; mood: rs > 0.99, *ps* < 0.001; intraclass coefficients > 0.99). Moreover, we computed cross-correlations between all generating (“generating”) and recovered (“fitted”) parameters. The resulting matrix showed high diagonal (choice winning model: rs > 0.37; mood winning model: rs > 0.99) and low off-diagonal (choice winning model: abs(rs) < 0.11; mood winning model: abs(rs) < 0.08) correlations, further supporting parameter recovery.

**Model fitting approach**: It is true that hierarchical Bayesian estimation and Markov chain Monte Carlo (MCMC) is a promising approach for model fitting (Ahn et al., 2017; Piray et al., 2019). However, the maximum likelihood-based model fitting procedure (MLE), e.g., fmincon in Matlab, were widely used, especially in domains of risky decision making and mood science (Blain & Rutledge, 2020; Rutledge, de Berker, et al., 2016; Rutledge et al., 2014), with the advantage of less time cost. Importantly, recently literature shows same pattern between two approaches (Rutledge et al., 2014; Vanhasbroeck et al., 2021). Specifically, Vanhasbroeck et al., (2021) reproduced all results in hierarchical way with that in MLE way in the context of gambling with momentary mood ratings (the same as the current task), suggesting reliability of MLE, at least for the current task. Excellent performance of parameter recovery and model recovery also demonstrated that MLE in this task was reliable.

**Parameter range:** The boundaries of parameter ranges were same as previous literature (Blain & Rutledge, 2020; Rutledge, de Berker, et al., 2016; Rutledge et al., 2014). These boundaries showed weakly-informative priors. Take an example of loss aversion ($\lambda$: 0.5-5). The value of $\lambda$ higher than 1 suggests loss aversion, while lower than 1 suggests loss seeking. The higher the $\lambda$ , the more loss aversion. This boundary allowed flexible feature for loss attitude. Based on the classic prospect theory, people, on average, are aversive to loss. Therefore, there was more space for loss aversion, while less space for loss seeking.

**R^2^:** Although R^2^ for continuous data or pseudoR^2^ for dichotomous data are important fitting matrix, e.g., assessment for fitting performance, limited research reported it. Based on the existing literature, pseudoR^2^ of 0.37 for the choice winning model was comparable to the previous literature using the same task: 0.46 (Rutledge, Smittenaar, et al., 2016), lower than that using the simple tasks (0.86 for effort-based decision-making task (Lockwood et al., 2021) and 0.54 for reinforcement learning task (Blain & Rutledge, 2020)), but higher than that using more complex tasks (pseudoR^2^ of 0.26 for two-step task (Daw et al., 2011)). For mood models, R^2^ of 0.42 was comparable to previous mood models (Blain & Rutledge, 2020; Rutledge, de Berker, et al., 2016; Rutledge et al., 2014).

**Supplementary Note 6: integration of mood into choice models**

Although we modeled choice and mood separately to examine cognitive and affective mechanisms underlying increased risk behavior in adolescent suicidal patients, one interesting question was whether mood responses influence subsequent gambling choices and how to model them. First, we median-split mood responses (except the final rating) to compare gambling rate. Results showed a trend for less gambling rate in higher mood (t = -1.971, p = 0.050). However, there was no significant group difference (F = 0.680, p = 0.507). Second, with the assumption that mood biases choice, we constructed mcM1 based on cM3 (the winning choice model).

$P_{gamble}= \frac{1-\beta_{val}}{1+e^{-\mu(U_{gamble}-U_{certain}+\beta_{Mood} zMood)}}+\beta_{val} if \beta_{val}\geq0$	(12)

$P_{gamble}= \frac{1+\beta_{val}}{1+e^{-\mu(U_{gamble}-U_{certain +\beta_{Mood} zMood})}} if \beta_{val}<0$	(13)

$\beta_{val}\left\{ \begin{aligned} \beta_{gain}, gain trials, \\ \beta_{loss}, loss trials. \end{aligned} \right.$	(14)

Based on our finding of the negative correlation between mood sensitivity to certain rewards and gambling rate in S^+^, we separated β_Mood_ parameter into β_Mood-CR_ and β_Mood-GR_ (cmM2).

$\beta_{Mood}\left\{ \begin{aligned} \beta_{Mood-CR}, certain choices, \\ \beta_{Mood-GR}, gamble choices. \end{aligned} \right.$	(15)

Model comparison using BIC supported cM3 (Table S7), that is, without consideration of mood in choice modeling. The mood bias parameters in neither cM2 nor cM3 reached significance (*ps* > 0.091), which may be due to the absence of a blocked design in our experiment, unlike in Vinckier et al. (2018) and Eldar and Niv (2015).

**Supplementary Note 7: other candidate models**

We also considered the traditional bias parameter (cM4), rather than approach/avoidance parameters. We limited the bias to the range of [-100, 100], which was in reward-equivalent units.

$P_{gamble}= \frac{1}{1+e^{-\mu(U_{gamble}-U_{certain}+\beta_{bias} )}}$	(16)

However, model comparison did not support cM4 (Table S7).

For the mood models, we additionally considered a term capturing whether participants gambled or not, independent of the gambling value (mM9), based on the winning model mM3. Given that the primary focus of the current study was the effect of STB, particularly in the S^+^ group, using the S^−^ and HC groups as baseline controls, we focused primarily on the winning model identified in the S^+^ group. Model comparison supported mM3 as the winning model in the S^+^ group (Table S8). Nevertheless, we also examined the results obtained with mM9. Overall, this additional analysis replicated the significant group differences in mood sensitivity to CR, both between S^+^ and S^−^ (t = -5.480, *p* = 0.015) and between S^+^ and HC (t = -2.025, *p* = 0.044), whereas S^−^ and HC did not differ significantly from each other (t = 0.628, *p* = 0.531). By contrast, no significant group differences were found in mood sensitivity to GR or in mood sensitivity to gambling (*ps* > 0.121).

**Supplementary Note 8: Clarification for FDR correction.**

In the clinical dataset we conducted a large number of inferential tests (χ², t-tests, ANOVAs, regressions) spanning: (i) group differences in demographic/clinical characteristics; (ii) sanity checks (e.g., anxiety/depression questionnaires); (iii) primary hypotheses (e.g., group differences in risky behavior); (iv) model-based analyses (parameter checks and between-group contrasts); and (v) control/sensitivity analyses. Post-hoc t-tests were performed only when the three-group ANOVA was significant. This yielded >150 p-values. FDR was applied using all these p-values.

**Supplementary Note 9: Mood model comparison using subjective values.**

To identify whether mood modeling was based on objective or subjective values, we constructed two model families: one in which mood was driven by objective monetary outcomes (objective values) and one in which mood was driven by subjective values derived from each participant’s fitted choice model (subjective values). We then used the VBA_groupBMC function in the VBA toolbox(Daunizeau et al., 2014) to perform family-wise model comparison, with 6 candidate mood models within each family. Consistent with previous literature, the objective-value family provided a clearly superior fit to the data (exceedance probability, EP = 1.000).

**References**

Ahn, W.-Y., Haines, N., & Zhang, L. (2017). Revealing Neurocomputational Mechanisms of Reinforcement Learning and Decision-Making With the hBayesDM Package. *Computational Psychiatry*, *1*(0), 24. https://doi.org/10.1162/CPSY_a_00002

Blain, B., & Rutledge, R. B. (2020). Momentary subjective well-being depends on learning and not reward. *ELife*, *9*. https://doi.org/10.7554/eLife.57977

Daunizeau, J., Adam, V., & Rigoux, L. (2014). VBA: A probabilistic treatment of nonlinear models for neurobiological and behavioural data. *PLoS Computational Biology*, *10*(1), e1003441. https://doi.org/10.1371/journal.pcbi.1003441

Daw, N. D., Gershman, S. J., Seymour, B., Dayan, P., & Dolan, R. J. (2011). Model-Based Influences on Humans’ Choices and Striatal Prediction Errors. *Neuron*, *69*(6), 1204–1215. https://doi.org/10.1016/j.neuron.2011.02.027

Eisenlohr-Moul, T. A., Miller, A. B., Giletta, M., Hastings, P. D., Rudolph, K. D., Nock, M. K., & Prinstein, M. J. (2018). HPA axis response and psychosocial stress as interactive predictors of suicidal ideation and behavior in adolescent females: a multilevel diathesis-stress framework. *Neuropsychopharmacology*, *43*(13), 2564–2571. https://doi.org/10.1038/s41386-018-0206-6

Glenn, C. R., Millner, A. J., Esposito, E. C., Porter, A. C., & Nock, M. K. (2019). Implicit Identification with Death Predicts Suicidal Thoughts and Behaviors in Adolescents. *Journal of Clinical Child & Adolescent Psychology*, *48*(2), 263–272. https://doi.org/10.1080/15374416.2018.1528548

Glenn, J. J., Werntz, A. J., Slama, S. J. K., Steinman, S. A., Teachman, B. A., & Nock, M. K. (2017). Suicide and self-injury-related implicit cognition: A large-scale examination and replication. *Journal of Abnormal Psychology*, *126*(2), 199–211. https://doi.org/10.1037/abn0000230

Goodfellow, B., Kõlves, K., & de Leo, D. (2018). Contemporary Nomenclatures of Suicidal Behaviors: A Systematic Literature Review. *Suicide and Life-Threatening Behavior*, *48*(3), 353–366. https://doi.org/10.1111/sltb.12354

Jollant, F., Colle, R., Nguyen, T. M. L., Corruble, E., Gardier, A. M., Walter, M., Abbar, M., & Wagner, G. (2023). Ketamine and esketamine in suicidal thoughts and behaviors: a systematic review. *Therapeutic Advances in Psychopharmacology*, *13*. https://doi.org/10.1177/20451253231151327

Lockwood, P. L., Abdurahman, A., Gabay, A. S., Drew, D., Tamm, M., Husain, M., & Apps, M. A. J. (2021). Aging Increases Prosocial Motivation for Effort. *Psychological Science*, *32*(5), 668–681. https://doi.org/10.1177/0956797620975781

Miller, A. B., Eisenlohr-Moul, T., Giletta, M., Hastings, P. D., Rudolph, K. D., Nock, M. K., & Prinstein, M. J. (2017). A within-person approach to risk for suicidal ideation and suicidal behavior: Examining the roles of depression, stress, and abuse exposure. *Journal of Consulting and Clinical Psychology*, *85*(7), 712–722. https://doi.org/10.1037/ccp0000210

Miller, A. B., Jenness, J. L., Elton, A. L., Pelletier-Baldelli, A., Patel, K., Bonar, A., Martin, S., Dichter, G., Giletta, M., Slavich, G. M., Rudolph, K. D., Hastings, P., Nock, M., Prinstein, M. J., & Sheridan, M. A. (2024). Neural Markers of Emotion Reactivity and Regulation Before and After a Targeted Social Rejection: Differences Among Girls With and Without Suicidal Ideation and Behavior Histories. *Biological Psychiatry*, *95*(12), 1100–1109. https://doi.org/10.1016/j.biopsych.2023.10.015

Millner, A. J., den Ouden, H. E. M., Gershman, S. J., Glenn, C. R., Kearns, J. C., Bornstein, A. M., Marx, B. P., Keane, T. M., & Nock, M. K. (2019). Suicidal thoughts and behaviors are associated with an increased decision-making bias for active responses to escape aversive states. *Journal of Abnormal Psychology*, *128*(2), 106–118. https://doi.org/10.1037/abn0000395

Neacsiu, A. D., Fang, C. M., Rodriguez, M., & Rosenthal, M. Z. (2018). Suicidal Behavior and Problems with Emotion Regulation. *Suicide and Life-Threatening Behavior*, *48*(1), 52–74. https://doi.org/10.1111/sltb.12335

O’Connor, R. C., & Nock, M. K. (2014). The psychology of suicidal behaviour. *The Lancet Psychiatry*, *1*(1), 73–85. https://doi.org/10.1016/S2215-0366(14)70222-6

Piray, P., Dezfouli, A., Heskes, T., Frank, M. J., & Daw, N. D. (2019). Hierarchical Bayesian inference for concurrent model fitting and comparison for group studies. *PLOS Computational Biology*, *15*(6), e1007043. https://doi.org/10.1371/journal.pcbi.1007043

Pompili, M., Amador, X. F., Girardi, P., Harkavy-Friedman, J., Harrow, M., Kaplan, K., Krausz, M., Lester, D., Meltzer, H. Y., Modestin, J., Montross, L. P., Bo Mortensen, P., Munk-Jørgensen, P., Nielsen, J., Nordentoft, M., Saarinen, P. I., Zisook, S., Wilson, S. T., & Tatarelli, R. (2007). Suicide risk in schizophrenia: learning from the past to change the future. *Annals of General Psychiatry*, *6*(1), 10. https://doi.org/10.1186/1744-859X-6-10

Poorolajal, J., Haghtalab, T., Farhadi, M., & Darvishi, N. (2016). Substance use disorder and risk of suicidal ideation, suicide attempt and suicide death: a meta-analysis. *Journal of Public Health*, *38*(3), e282–e291. https://doi.org/10.1093/pubmed/fdv148

Rutledge, R. B., de Berker, A. O., Espenhahn, S., Dayan, P., & Dolan, R. J. (2016). The social contingency of momentary subjective well-being. *Nature Communications*, *7*(1), 11825. https://doi.org/10.1038/ncomms11825

Rutledge, R. B., Skandali, N., Dayan, P., & Dolan, R. J. (2014). A computational and neural model of momentary subjective well-being. *Proceedings of the National Academy of Sciences*, *111*(33), 12252–12257. https://doi.org/10.1073/pnas.1407535111

Rutledge, R. B., Smittenaar, P., Zeidman, P., Brown, H. R., Adams, R. A., Lindenberger, U., Dayan, P., & Dolan, R. J. (2016). Risk Taking for Potential Reward Decreases across the Lifespan. *Current Biology*, *26*(12), 1634–1639. https://doi.org/10.1016/j.cub.2016.05.017

Sarchiapone, M., Carli, V., Cuomo, C., & Roy, A. (2007). Childhood trauma and suicide attempts in patients with unipolar depression. *Depression and Anxiety*, *24*(4), 268–272. https://doi.org/10.1002/da.20243

Tsypes, A., Hallquist, M. N., Ianni, A., Kaurin, A., Wright, A. G. C., & Dombrovski, A. Y. (2024). Exploration-Exploitation and Suicidal Behavior in Borderline Personality Disorder and Depression. *JAMA Psychiatry*, *81*(10), 1010. https://doi.org/10.1001/jamapsychiatry.2024.1796

Vanhasbroeck, N., Devos, L., Pessers, S., Kuppens, P., Vanpaemel, W., Moors, A., & Tuerlinckx, F. (2021). Testing a computational model of subjective well-being: a preregistered replication of Rutledge et al. (2014). *Cognition and Emotion*, *35*(4), 822–835. https://doi.org/10.1080/02699931.2021.1891863

Wang, Z., Wang, T., Nan, T., Xu, J., Aleman, A., Luo, Y., Blain, B., Liu, Y., & Xu, P. (2025). *Dissociable Roles of Reward Prediction Error in the Contrasting Mood Dynamics of Depression and Anxiety*. https://doi.org/10.31234/osf.io/z6s2r_v1

**Figure S1.** Suicidal Attempts vs. Suicidal Thoughts.

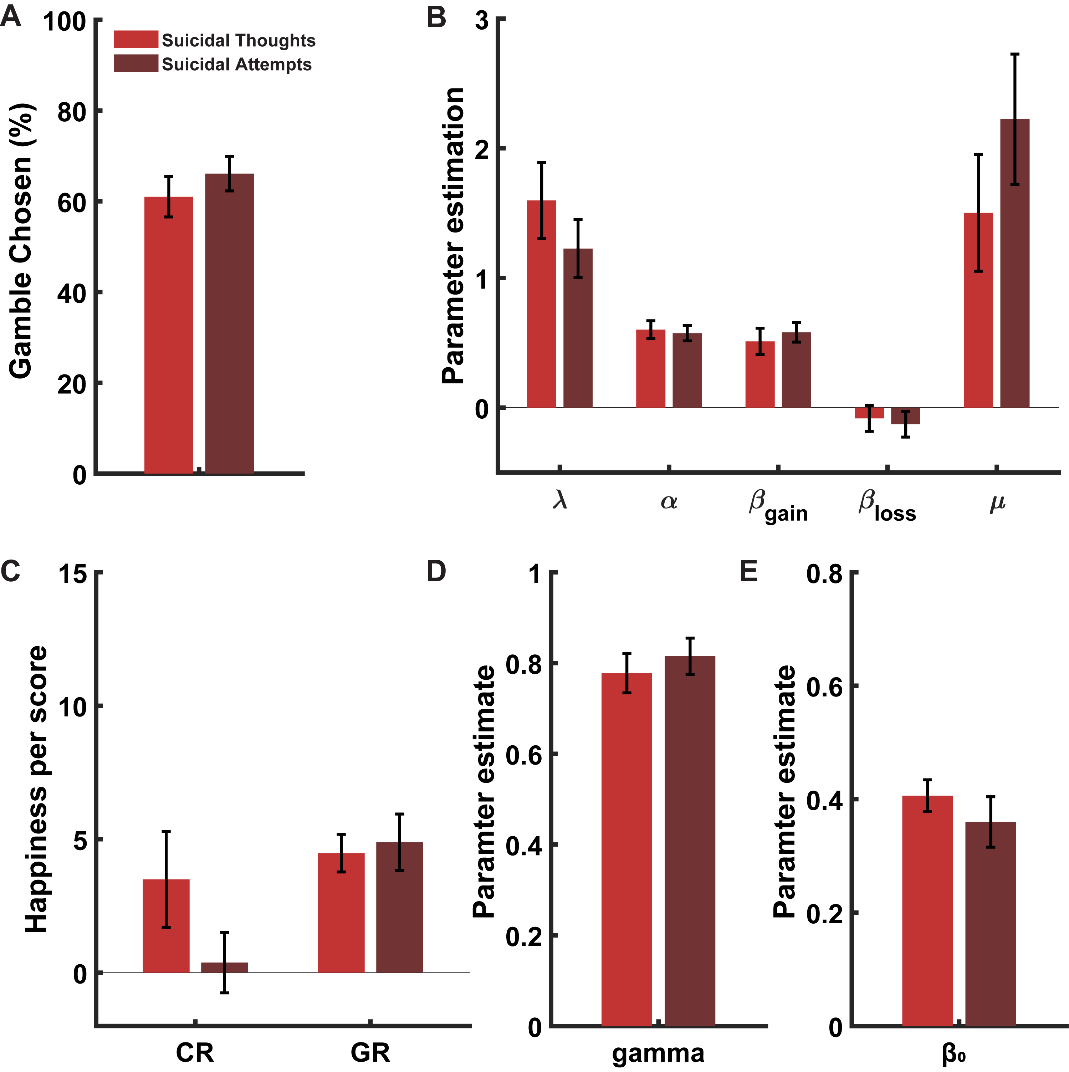

**Figure S2.** Control analysis for age and other anxiolytics in patients, which were significant between groups.

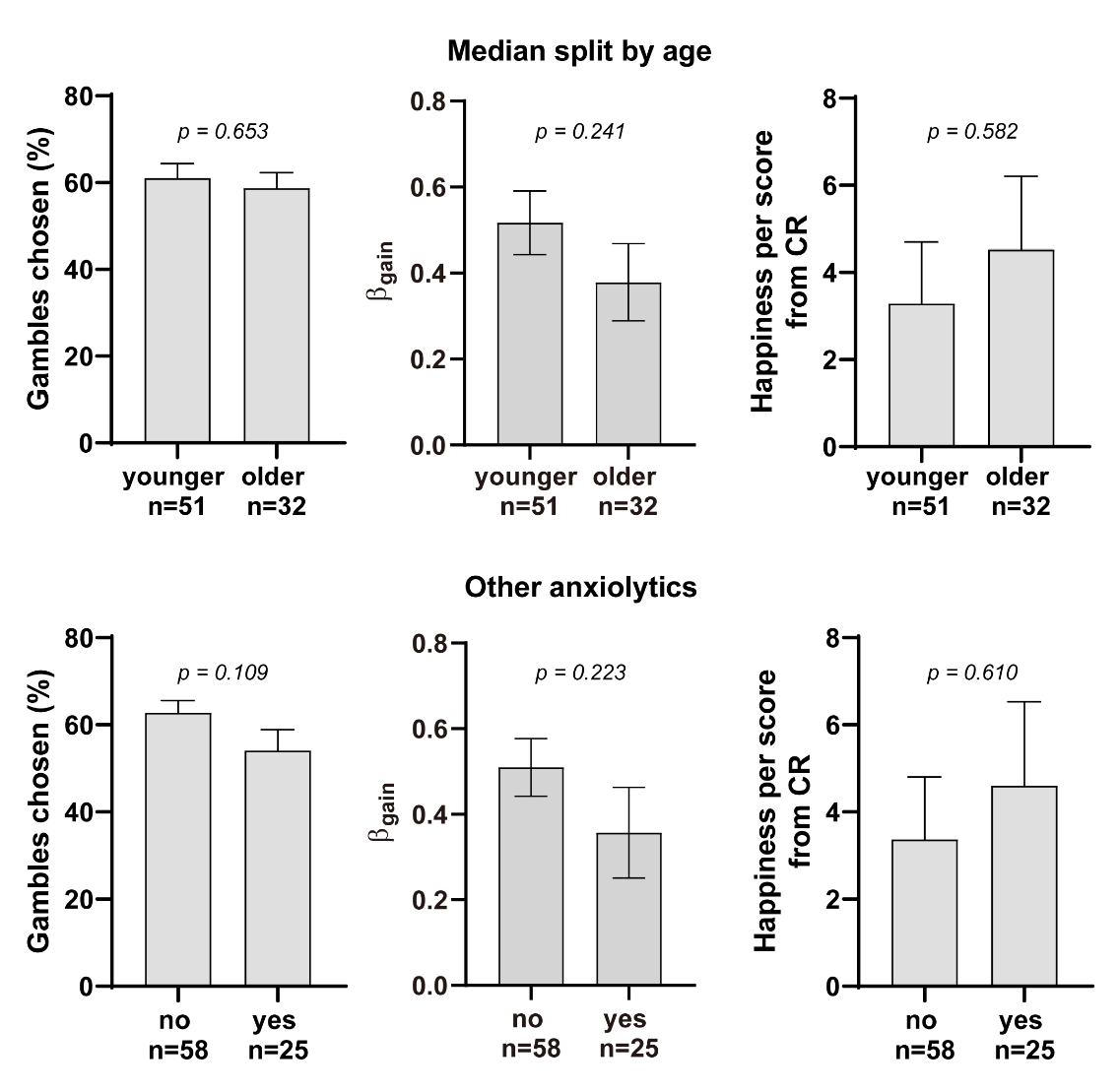

**Figure S3.** Control analysis for childhood maltreatment and emotion regulation problems in patients, which were significant between groups.

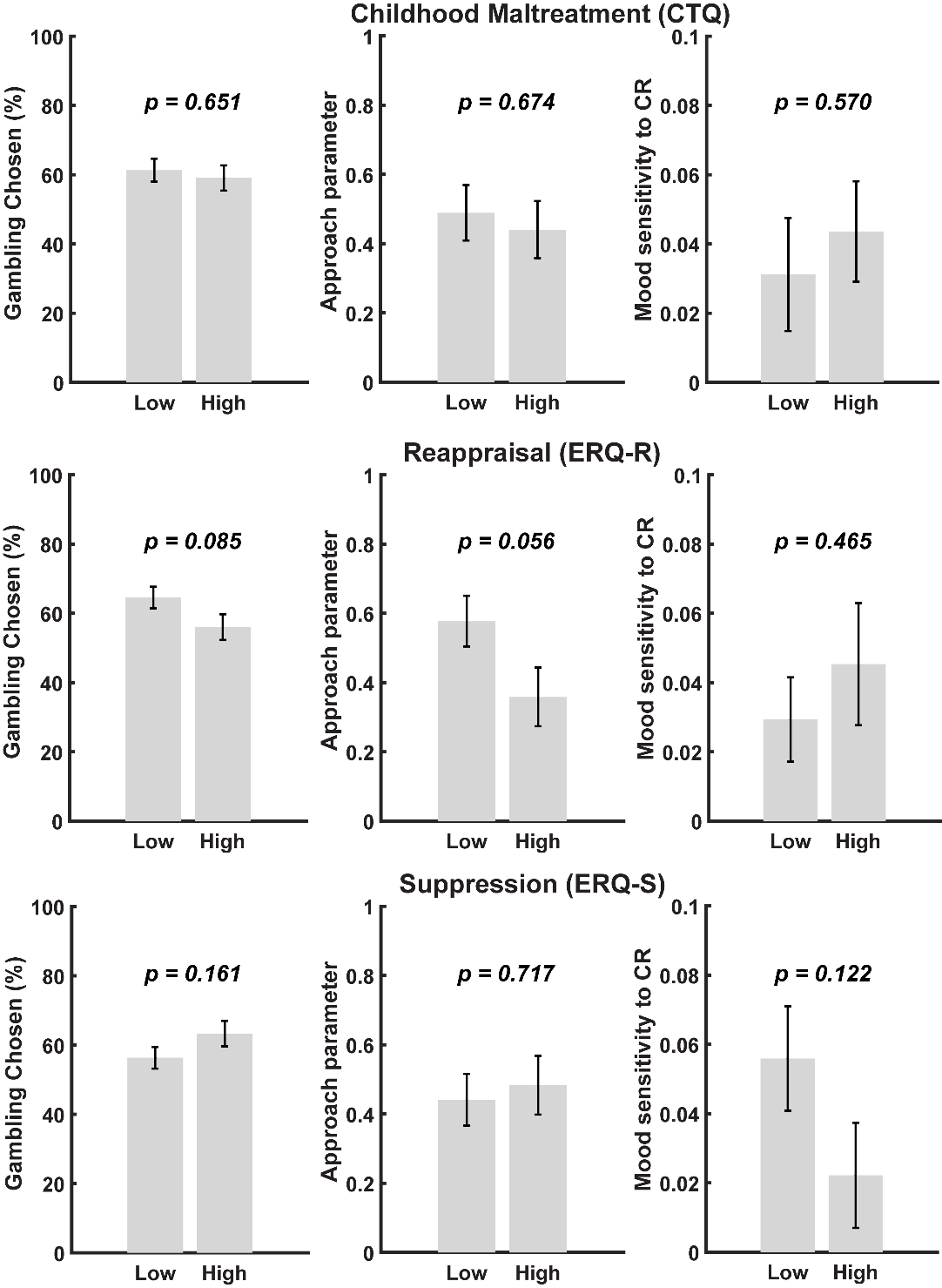

**Figure S4.** Control analysis for depression/anxiety symptoms in patients, which were significant between groups.

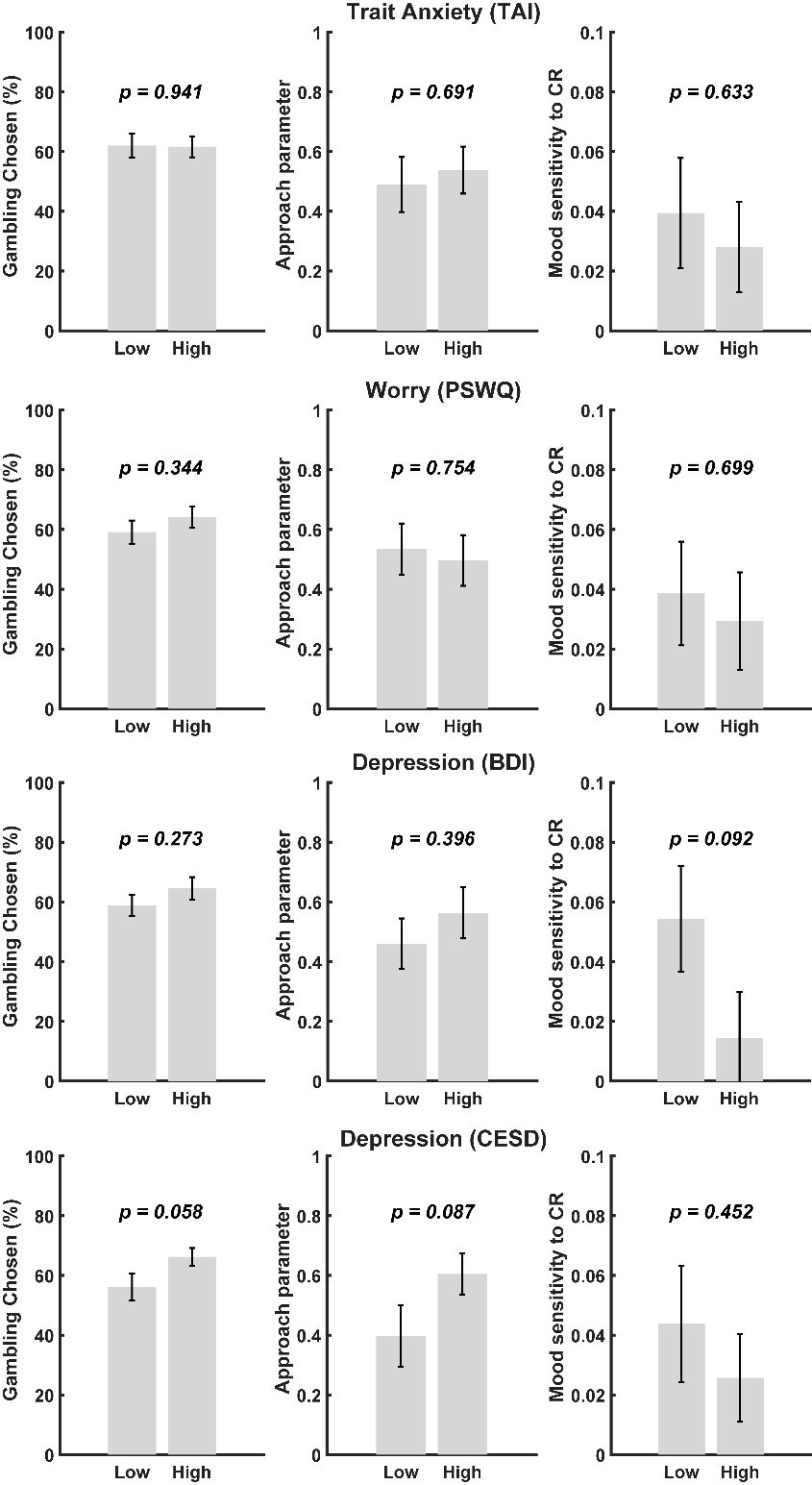

**Figure S5.** Choice and mood model recovery. Mood model recovery was performed in a stage-by-stage manner, mirroring the model comparison procedure used in the main analyses because each model comparison step addresses a specific question and doing so prevents dispersion of model evidence across multiple similar models. Specifically, rather than entering all mood models into a single recovery space, recovery was evaluated separately within each comparison space used for model selection. Thus, each confusion matrix reflects recovery among only the models that were directly compared at that stage.

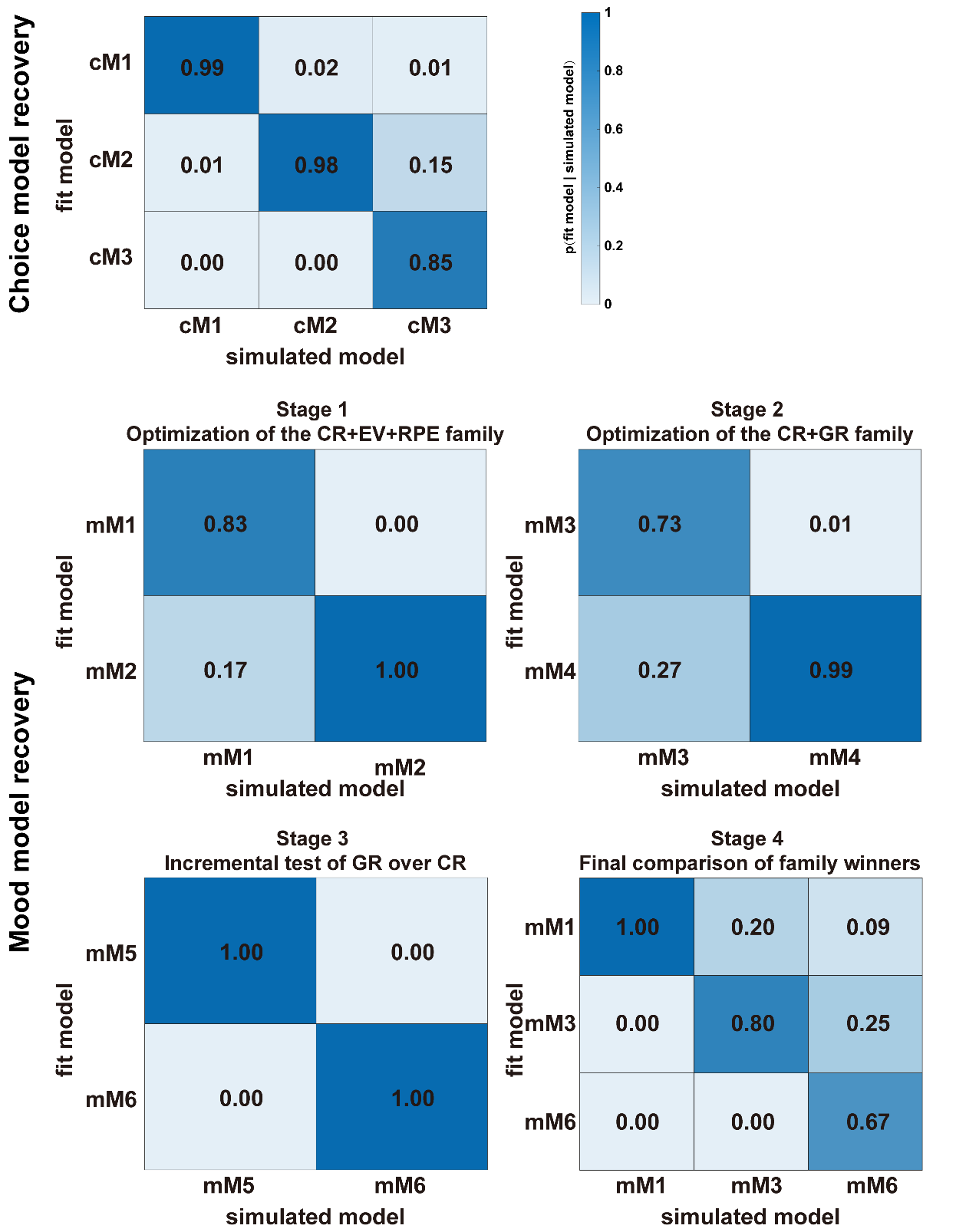

**Figure S6.** Parameter recovery for the winning choice and mood models (cM3; mM3).

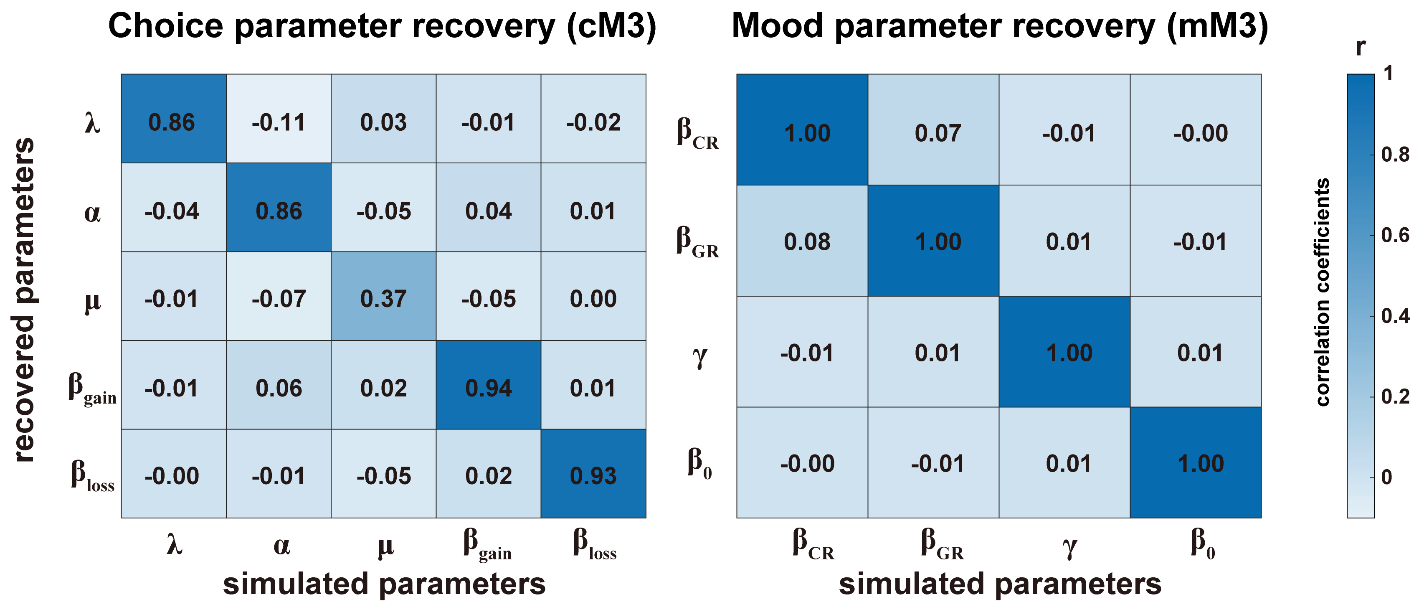

**Figure S7.** Replication of Rutledge et.al., (2017)’s findings using BDI. Depression symptom measured by BDI was negatively correlated with the baseline mood parameter.

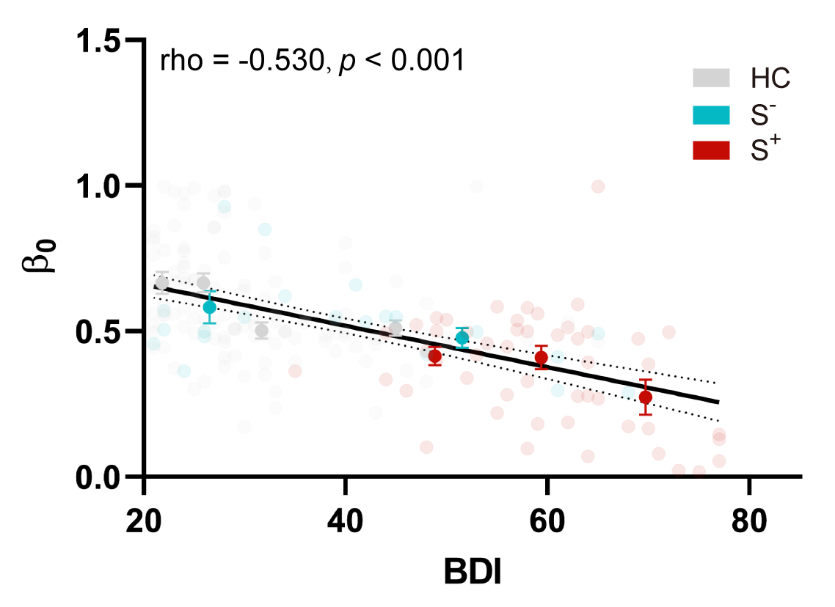

**Figure S8.** RPE model results (M1). A) Group differences in mood sensitivity to certain reward (CR), expected value (EV), and reward prediction error (RPE). B) Correlation between Suicidal Ideation score at current time (BSI-C) and mood sensitivity to CR. Abbreviations: HC, healthy control; S^-^, patients without suicidal thoughts and behavior; S^+^, patients with suicidal thoughts and behavior; BSI-C, Beck Scale for Suicidal Ideation at the current time; **p*<0.05.

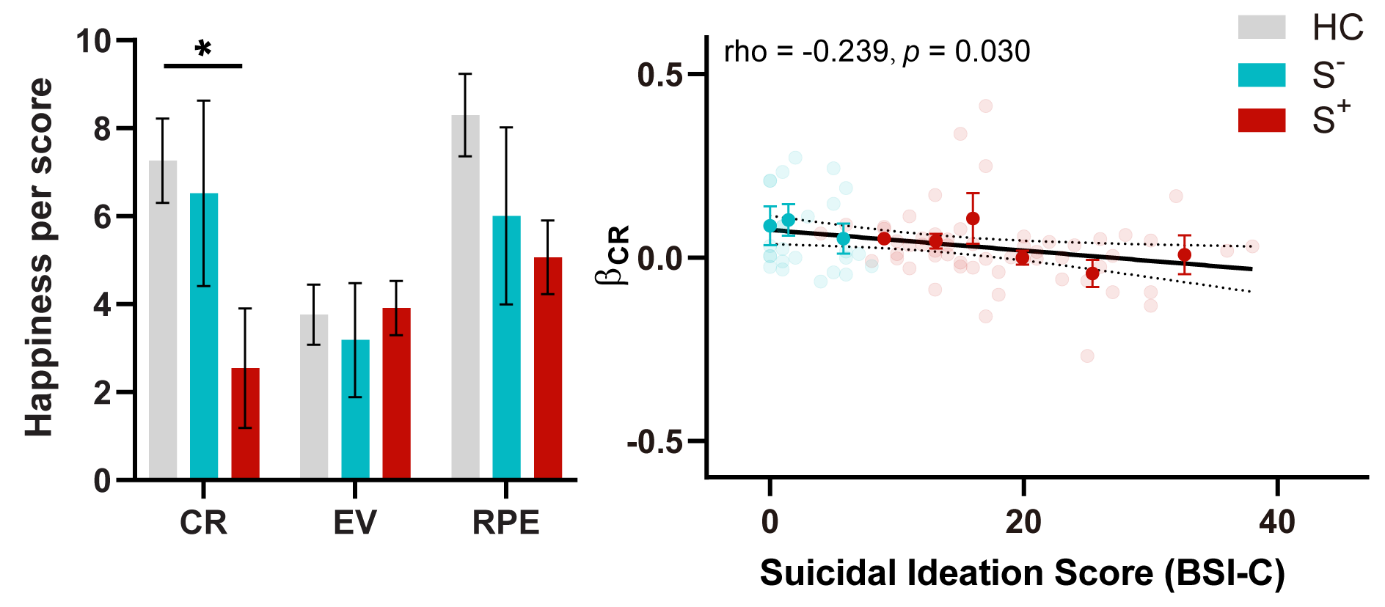

**Figure S9.** Expectation effect on mood. A) Group differences in mood sensitivity to certain reward (CR), gamble reward (GR), and expected value (EV). B) Correlation between Suicidal Ideation score at current time (BSI-C) and mood sensitivity to CR. Abbreviations: HC, healthy control; S^-^, patients without suicidal thoughts and behavior; S^+^, patients with suicidal thoughts and behavior; BSI-C, Beck Scale for Suicidal Ideation at the current time; **p*<0.05.

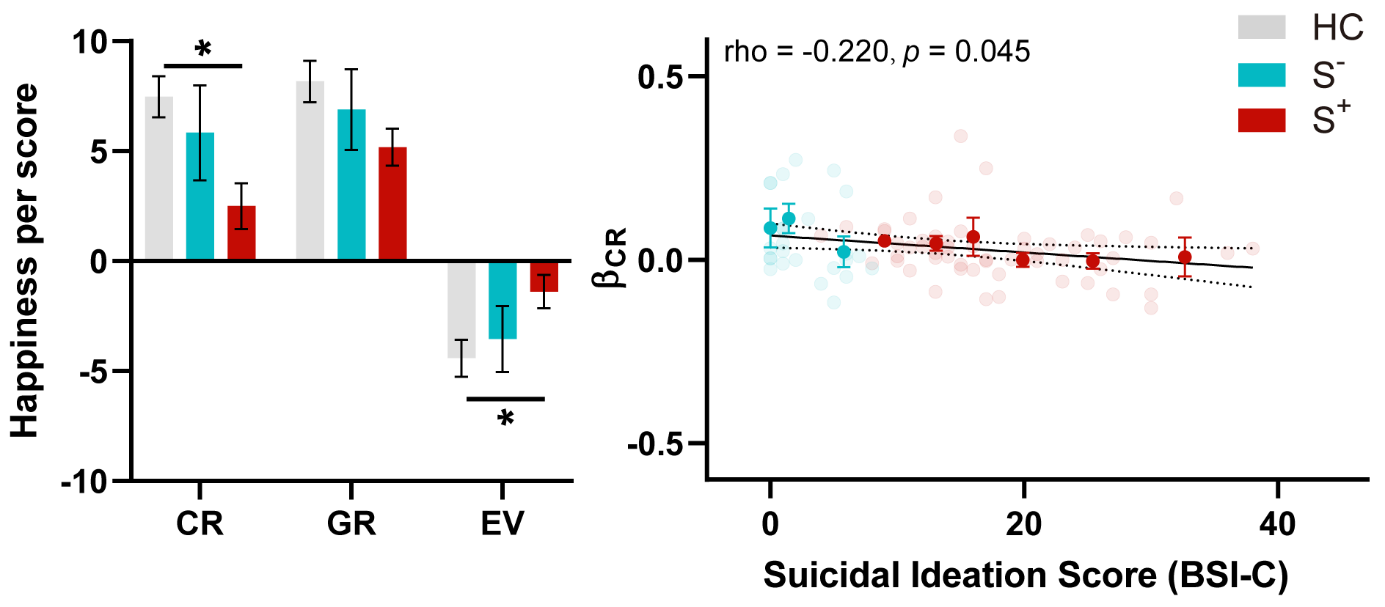

**Figure S10.** Results from M5. A) Group differences in mood sensitivity to certain reward (CR), better gamble reward (GRbetter), and worse gamble reward (GRworse). B) Correlation between Suicidal Ideation score at current time (BSI-C) and mood sensitivity to CR. Abbreviations: HC, healthy control; S^-^, patients without suicidal thoughts and behavior; S^+^, patients with suicidal thoughts and behavior; BSI-C, Beck Scale for Suicidal Ideation at the current time; **p*<0.05.

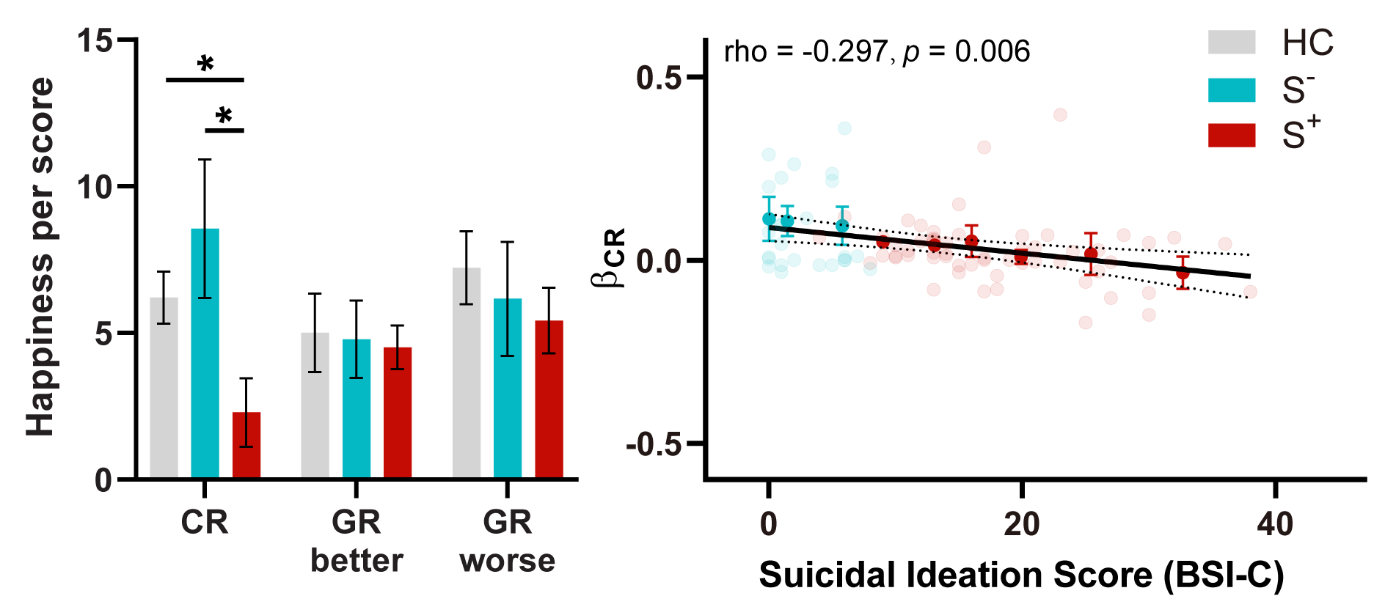

**Figure S11.** Permutation tests. We conducted permutation tests (1,000,000 iterations) to evaluate the robustness of our main results, ensuring consistency with the same normal distribution and sample size. Specifically, in each permutation, we randomly drew five samples from the S^+^ group and repeated this process 20 times to construct a suicidal group (100 samples). The same procedure was applied to the S^−^ group to construct a control group. We then calculated the t-values between the two constructed groups on variables of interest, including the proportion of gambling choices, the approach parameter, and mood sensitivity to certain rewards (CR). The 1,000,000 t-values formed the H1 distributions of between-group differences. Nonparametric P-values were calculated as the proportion of permutations that generated t-values failing to reach significance (parametric p>0.05), divided by 1,000,000. Across these variables, the S^+^ group significantly differed from the S^−^ group (Ps < 0.038).

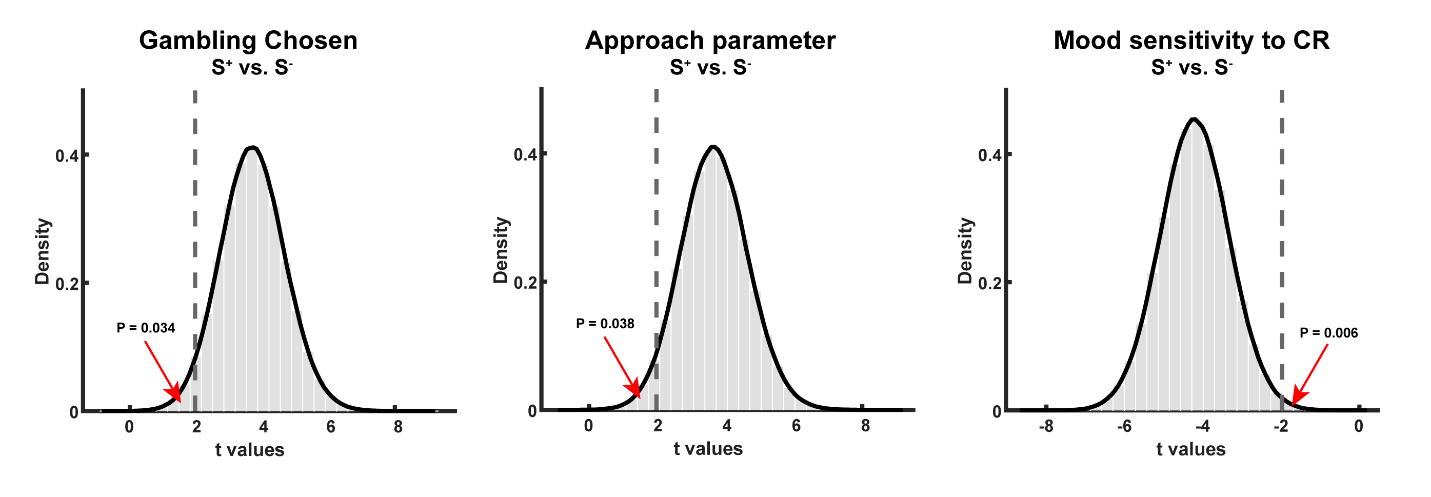

**Table S1**: A short summary for risk measurement in STB. Abbreviations: BIS, Barratt Impulsiveness Scale; IGT, Iowa Gambling Task; CGT, Cambridge Gambling Task; BART, the Balloon analog risk task.

| **Study** | **Measurements** | **Tools** | **Analysis level** | **Hypotheses** | **Results** | **Category** | **Model specification** |
| --- | --- | --- | --- | --- | --- | --- | --- |
| Millner et al., 2020 | Questionnaire | UPPS-P Impulsive Behavior Scale +BIS | Sum (sub)scale scores | Heightened impulsiveness in STB | ns | Self-report |  |
| Zakowicz et al., 2021 | Questionnaire | BIS | Sum (sub)scale scores | Heightened impulsiveness in STB | ns | Self-report |  |
| Jollant et al., 2005 | Task | IGT | Model-agnostic | More risky behavior in STB | Task performance: STB<control | Risk+Ambiguity+Learning |  |
| Bridge et al., 2012 | Task | IGT | Model-agnostic | More risky behavior in STB | Task performance: STB<control | Risk+Ambiguity+Learning |  |
| Martino et al., 2010 | Task | IGT | Model-agnostic | More risky behavior in STB | Task performance: STB<control | Risk+Ambiguity+Learning |  |
| Chamberlain et al., 2013 | Task | CGT | Model-agnostic | More risky (irrational) behavior in STB | Proportion of rational choices: STB<control | Risk |  |
| Ackerman et al., 2015 | Task | CGT | Model-agnostic | More risky behavior in STB | Proportion of bet: STB>control | Risk |  |
| Dir et al., 2020 | Task | BART | Model-agnostic | More risky behavior in STB | Task performance: STB<control | Risk+Ambiguity+Learning |  |
| Liu et al., 2022 | Task | BART | Model-based | Decision-making bias in STB | Task performance: STB>control  Loss aversion: STB>control | Risk+Ambiguity+Learning | Exponential‐Weight Model: loss aversion, risk preference, updating exponent, prior belief of exploding |
| Baek et al., 2016-risk | Task | Gambling (gain + loss) | Model-based | Heightened risk aversion in STB | risk aversion: STB>control | Risk | Risk discount model: discount parameter |
| Baek et al., 2016-loss | Task | Gambling (mix) | Model-based | Heightened loss aversion in STB | loss aversion: STB>control | risk | Psychophysics;  indifference point |
| Alacreu-Crespo et al., 2020 | Task | IGT | Model-based | More risky behavior in STB | Task performance: STB<control;  Loss aversion: STB<control;  Learning: STB>control | Risk+Ambiguity+Learning | Prospect valence learning delta model: learning/memory, choice consistency, loss aversion, and feedback sensibility |
| The current study | Task | Gambling (gain + loss + mix) | Model-based | More risky behavior in STB | Gambling behavior: STB>control;  Approach parameter: STB>control | Risk | The Approach-Avoidance Prospect Theory Model: risk aversion, loss aversion, approach motivation, avoidance motivation, decision noise |

**Table S2**. Contrasts for demographic and psychological characteristics.

|  | **S^-^ vs. HC** | |  | **S^+^ vs. HC** | |
| --- | --- | --- | --- | --- | --- |
|  | **t/χ^2^** | ***p*** |  | **t/χ^2^** | ***p*** |
| Gender | 0.002 | 0.967 |  | 0.880 | 0.348 |
| Age | 0.816 | 0.416 |  | -1.458 | 0.147 |
| BSI-C | 2.030 | 0.044 |  | 19.889 | <0.001 |
| BSI-W | 0.342 | 0.733 |  | 23.723 | <0.001 |
| CTQ | 3.426 | <0.001 |  | 8.822 | <0.001 |
| ERQ-R | -1.609 | 0.110 |  | -8.278 | <0.001 |
| ERQ-S | 2.409 | 0.017 |  | 6.650 | <0.001 |
| TAI | 3.174 | 0.002 |  | 15.653 | <0.001 |
| PSWQ | 2.142 | 0.034 |  | 13.364 | <0.001 |
| BDI | 3.299 | 0.001 |  | 17.368 | <0.001 |
| CESD | 3.522 | <0.001 |  | 16.255 | <0.001 |

**Table S3**. Sample size for each diagnosis with and without comorbidity in S+ and S- groups.

| **Diagnosis** | **S^+^** | **S^-^** | **Statistics** |
| --- | --- | --- | --- |
| GAD | 1 | 2 | χ^2^ = 4.843  *p* = 0.304 |
| MDD | 25 | 9 |  |
| BD | 7 | 6 |  |
| MDD &BD | 2 | 0 |  |
| MDD & AD | 23 | 8 |  |

**Table S4**. Bivariate correlations between choice parameters and socio-demographic clinical variables. All *p* values were above 0.05.

|  |  | **λ** | **α** | **β_gain_** | **β_loss_** | **μ** |
| --- | --- | --- | --- | --- | --- | --- |
| **Demographics** | **Gender** | rho = 0.00,  *p* = 1.000 | rho = 0.15,  *p* = 0.179 | rho = 0.08,  *p* = 0.495 | rho = 0.14,  *p* = 0.197 | rho = -0.00,  *p* = 0.961 |
|  | **Age** | rho = 0.13,  *p* = 0.230 | rho = 0.07,  *p* = 0.517 | rho = -0.19,  *p* = 0.079 | rho = -0.03,  *p* = 0.811 | rho = -0.17,  *p* = 0.114 |
| **Social variables** | **CTQ** | rho = 0.01,  *p* = 0.946 | rho = -0.19,  *p* = 0.090 | rho = 0.08,  *p* = 0.495 | rho = 0.04,  *p* = 0.700 | rho = 0.20,  *p* = 0.075 |
|  | **ERQ-E** | rho = 0.14,  *p* = 0.215 | rho = -0.11,  *p* = 0.338 | rho = -0.17,  *p* = 0.114 | rho = -0.21,  *p* = 0.059 | rho = -0.03,  *p* = 0.769 |
|  | **ERQ-S** | rho = -0.13,  *p* = 0.230 | rho = -0.04,  *p* = 0.727 | rho = 0.15,  *p* = 0.187 | rho = 0.10,  *p* = 0.346 | rho = 0.17,  *p* = 0.128 |
| **Clinical variables** | **Illness duration** | rho = 0.08,  *p* = 0.480 | rho = -0.04,  *p* = 0.737 | rho = -0.05,  *p* = 0.650 | rho = -0.02,  *p* = 0.849 | rho = -0.10,  *p* = 0.351 |
|  | **Family history** | rho = 0.16,  *p* = 0.142 | rho = 0.04,  *p* = 0.706 | rho = 0.05,  *p* = 0.648 | rho = -0.04,  *p* = 0.700 | rho = -0.01,  *p* = 0.908 |
|  | **MDD** | rho = -0.11,  *p* = 0.317 | rho = 0.17,  *p* = 0.123 | rho = 0.02,  *p* = 0.882 | rho = -0.04,  *p* = 0.697 | rho = -0.11,  *p* = 0.324 |
|  | **GAD** | rho = 0.11,  *p* = 0.335 | rho = 0.16,  *p* = 0.162 | rho = 0.02,  *p* = 0.851 | rho = 0.03,  *p* = 0.797 | rho = -0.03,  *p* = 0.755 |
|  | **BD** | rho = 0.17,  *p* = 0.124 | rho = -0.12,  *p* = 0.288 | rho = 0.03,  *p* = 0.801 | rho = 0.04,  *p* = 0.707 | rho = 0.05,  *p* = 0.630 |

**Table S5**. Bivariate correlations between mood parameters and socio-demographic clinical variables. *P* values lower than 0.05 was highlighted in bold.

|  |  | **β_CR_** | **β_GR_** | **γ** | **β_o_** |
| --- | --- | --- | --- | --- | --- |
| **Demographics** | **Gender** | rho = 0.15,  *p* = 0.168 | rho = 0.23,  *p* = 0.033 | rho = 0.08,  *p* = 0.465 | rho = -0.17,  *p* = 0.126 |
|  | **Age** | rho = 0.07,  *p* = 0.503 | **rho = -0.27,**  ***p* = 0.012** | rho = 0.06,  *p* = 0.615 | **rho = 0.28,**  ***p* = 0.010** |
| **Social variables** | **CTQ** | rho = 0.04,  *p* = 0.739 | rho = 0.07,  *p* = 0.545 | rho = -0.06,  *p* = 0.610 | rho = -0.02,  *p* = 0.866 |
|  | **ERQ-E** | rho = 0.09,  *p* = 0.440 | **rho = -0.26,**  ***p* = 0.017** | rho = -0.13,  *p* = 0.255 | **rho = 0.46,**  ***p* < 0.001** |
|  | **ERQ-S** | rho = -0.19,  *p* = 0. 086 | rho = -0.06,  *p* = 0.599 | rho = -0.14,  *p* = 0.217 | rho = -0.16,  *p* = 0.142 |
| **Clinical variables** | **Illness duration** | rho = -0.01,  *p* = 0.910 | rho = -0.10,  *p* = 0.366 | rho = -0.03,  *p* = 0.788 | rho = -0.02,  *p* = 0.861 |
|  | **Family history** | rho = 0.06,  *p* = 0.590 | rho = -0.01,  *p* = 0.908 | rho = -0.09,  *p* = 0.403 | rho = 0.13,  *p* = 0.231 |
|  | **MDD** | rho = -0.02,  *p* = 0.864 | rho = 0.09,  *p* = 0.396 | rho = -0.07,  *p* = 0.521 | rho = -0.21,  *p* = 0.053 |
|  | **GAD** | rho = -0.06,  *p* = 0.569 | rho = 0.05,  *p* = 0.627 | rho = 0.01,  *p* = 0.912 | rho = -0.08,  *p* = 0.491 |
|  | **BD** | rho = 0.16,  *p* = 0.153 | rho = -0.07,  *p* = 0.542 | rho = -0.02,  *p* = 0.888 | rho = 0.16,  *p* = 0.160 |

**Table S6.** Mood model comparison by separating gambling outcomes into better and worse parts.

| Model # | Model specification | # of parameters | Δ BIC | meanR^2^ | Δ BIC for each group | | |
| --- | --- | --- | --- | --- | --- | --- | --- |
|  |  |  |  |  | HC | S^-^ | S^+^ |
| **mM3** | **β_0_, β_CR_, β_GR_, γ** | **4** | **0** | **0.42** | **0** | **0** | **0** |
| mM7 | β_0_, β_CR_, β_GR_better_, β_GR_worse_, γ | 5 | **-331.48** | 0.49 | **-355.10** | **-6.56** | 30.18 |
| mM8 | β_0_, β_CR_, β_GR_better_, β_GR_worse_, γ_CR_, γ_GR_better_, γ_GR_better_ | 7 | -105.40 | 0.56 | -313.09 | 81.34 | 126.34 |

Abbreviations: Δ BIC, Bayesian information criterion relative to the winning model in S^+^ group (mM3); HC, healthy control; S^-^, patients without suicidal thoughts and behavior; S^+^, patients with suicidal thoughts and behavior.

**Table S7.** Choice model comparison by integrating mood or adding traditional bias.

| Model # | Model specification | # of parameters | Δ BIC | meanR^2^ | Δ BIC for each group | | |
| --- | --- | --- | --- | --- | --- | --- | --- |
|  |  |  |  |  | HC | S- | S+ |
| **cM3** | **λ, α, β_gain_, β_loss_, µ** | **5** | **0** | **0.37** | **0** | **0** | **0** |
| cmM1 | λ, α, β_gain_, β_loss_, µ, β_Mood_ | 6 | 5530.23 | 0.39 | 287.20 | 68.76 | 174.27 |
| cmM2 | λ, α, β_gain_, β_loss_, µ, β_Mood-CR,_ β_Mood-GR_ | 7 | 1031.18 | 0.41 | 577.03 | 123.84 | 330.32 |
| cM4 | **λ, α, µ, β_bias_** | 4 | 376.75 | 0.32 | 277.65 | 34.31 | 64.79 |

Abbreviations: Δ BIC, Bayesian information criterion relative to the winning model (cM3); HC, healthy control; S^-^, patients without suicidal thoughts and behavior; S^+^, patients with suicidal thoughts and behavior.

**Table S8.** Mood model comparison by adding a term for whether participants gambled or not, independent of the gambling value.

| Model # | Model specification | # of parameters | Δ BIC | meanR^2^ | Δ BIC for each group | | |
| --- | --- | --- | --- | --- | --- | --- | --- |
|  |  |  |  |  | HC | S^-^ | S^+^ |
| **mM3** | **β_0_, β_CR_, β_GR_, γ** | **4** | **0** | **0.42** | **0** | **0** | **0** |
| mM9 | β_0_, β_CR_, β_GR_ β_gamble_, γ | 5 | **-373.13** | 0.49 | **-371.83** | **-40.74** | 39.45 |

Abbreviations: Δ BIC, Bayesian information criterion relative to the winning model in S^+^ group (mM3); HC, healthy control; S^-^, patients without suicidal thoughts and behavior; S^+^, patients with suicidal thoughts and behavior.

**Table S9.** Bayesian independent sample t-tests of median-split anxiety and depression scores (including TAI, PSWQ, BDI, and CESD) on main results (gambling rate, approach parameter (β_gain_), and mood sensitivity to certain rewards (**β_CR_**)) support that general symptoms of anxiety and depression overall did not influence our main results. BF₀₁ is a Bayes factor comparing the null model (M₀) to the alternative model (M₁), where M₀ assumes no group difference. BF₀₁ > 1 indicates that evidence favors M₀. Generally, Bayes Factors between 1 and 3 were interpreted as anecdotal evidence and between 3 and 10 as moderate evidence.

| **BF_01_** | **TAI** | **PSWQ** | **BDI** | **CESD** |
| --- | --- | --- | --- | --- |
| **Gambling rate** | 4.080 | 2.768 | 2.425 | 0.820 |
| **β_gain_** | 3.819 | 3.911 | 2.987 | 1.128 |
| **β_CR_** | 3.704 | 3.826 | 1.185 | 3.178 |

**Table S10.** Linear regressions of gambling behaviour, value-insensitive approach parameter (**β_gain_**), and mood sensitivity to certain rewards (**β_CR_**) on group as a predictor (1 for S^+^ group and 0 for S^-^ group) and scores for anxiety and depression as covariates.

|  | **Gambling rate** | **β_gain_** | **β_CR_** |
| --- | --- | --- | --- |
| **Group** | **β = 0.164, t = 2.305,**  ***p* = 0.024** | **β = 0.374, t = 2.257,**  ***p* = 0.027** | **β = -0.105, t = -3.461,**  ***p* = 0.001** |
| **TAI** | β = -0.010, t = -1.796,  *p* = 0.077 | β = -0.008, t = -0.649,  *p* = 0.519 | β = 0.005, t = 1.921,  *p* = 0.059 |
| **PSWQ** | β = -0.001, t = -0.401,  *p* = 0.690 | β = -0.008, t = -0.968,  *p* = 0.337 | β = -0.002, t = -1.282,  *p* = 0.204 |
| **BDI** | β = 0.002, t = 0.519,  *p* = 0.606 | β = -0.003, t = -0.272,  *p* = 0.787 | **β = -0.004, t = -2.302,**  ***p* = 0.025** |
| **CESD** | β = 0.006, t = 1.585,  *p* = 0.118 | β = 0.015, t = 1.652,  *p* = 0.103 | β = 0.003, t = 1.942,  *p* = 0.056 |

**Table S11.** Linear regressions of gambling behaviour, value-insensitive approach parameter (**β_gain_**), and mood sensitivity to certain rewards (**β_CR_**) on group as a predictor (1 for S^+^ group and 0 for S^-^ group) and orthogonal components of anxiety and depression as covariates.

|  | **Gambling rate** | **β_gain_** | **β_CR_** |
| --- | --- | --- | --- |
| **Group** | **β = 0.164, t = 2.305,**  ***p* = 0.024** | **β = 0.378, t = 2.257,**  ***p* = 0.027** | **β = -0.105, t = -3.461,**  ***p* = 0.001** |
| **PC1** | β = -0.007, t = -0.425,  *p* = 0.672 | β = -0.010, t = -0.247,  *p* = 0.806 | β = 0.009, t = 1.169,  *p* = 0.247 |
| **PC2** | β = -0.073, t = -1.569,  *p* = 0.122 | β = -0.164, t = -1.509,  *p* = 0.136 | β = -0.011, t = -0.532,  *p* = 0.596 |
| **PC3** | β = 0.098, t = 1.429,  *p* = 0.158 | β = 0.205, t = 1.283,  *p* = 0.204 | β = 0.037, t = 1.255,  *p* = 0.214 |
| **PC4** | β = -0.086, t = -1.133,  *p* = 0.262 | β = 0.010, t = 0.054,  *p* = 0.957 | **β = 0.086, t = 2.657,**  ***p* = 0.010** |
